# HLA-Resolve: Four-field HLA typing and MHC variant detection from long-read hybrid capture

**DOI:** 10.64898/2026.03.27.26349549

**Authors:** Matthew R. Glasenapp, Muh-Ching Yee, Alexander E. Symons, Jeffrey G. Sheh, Obed A. Garcia, Omar E. Cornejo

## Abstract

The major histocompatibility complex (MHC) is among the most polymorphic and difficult-to-genotype regions of the human genome. Accurate typing of its HLA genes, which encode antigen-presenting molecules, is critical for transplantation, pharmacogenomics, and disease-risk prediction. Long-read sequencing is increasingly used in HLA typing because of its ability to phase variants across long distances, but many long-read HLA typing workflows still rely on long-range PCR. Here, we present an end-to-end workflow for MHC genotyping that pairs hybrid capture with long-read sequencing on PacBio or Oxford Nanopore, using single-step enzymatic fragmentation and barcoding to enable automated library preparation. The hybrid capture panel enriches both classical and non-classical HLA Class I and Class II genes, as well as all 69 protein-coding MHC Class III genes. We introduce HLA-Resolve, a bioinformatic tool that types HLA genes from PacBio reads via phased full-gene sequence reconstruction, and we benchmark the capture assay and HLA-Resolve together across 32 geographically diverse samples. With PacBio data, variant calling achieved F1 scores of 99.8% for SNVs and 98.4% for indels against the Genome in a Bottle benchmark. HLA-Resolve showed 99.5% allele concordance with reference typings from the International Histocompatibility Working Group and the Human Pangenome Reference Consortium (HPRC) at three-field resolution and 90.5% at four-field resolution, outperforming three other open-source long-read HLA typers on our HiFi hybrid capture reads. The full-gene sequences reconstructed by HLA-Resolve showed zero edit distance to the corresponding HPRC assemblies in 95% of comparisons. HLA-Resolve also showed high accuracy with whole-genome sequencing (WGS) data, achieving 99.4% two-field concordance across 146 public PacBio WGS libraries and 99.5% three-field concordance across the 52 with full-resolution reference typings. Although developed for the MHC, the workflow is customizable to any gene set, providing a general framework for high-resolution genotyping.

## Main

The major histocompatibility complex (MHC; 6p21.3) contains key genes involved in immune recognition and response, shaping host-pathogen interactions, susceptibility to infectious and autoimmune diseases, and donor-recipient compatibility in transplantation^1^. It is among the most polymorphic regions of the genome^2–4^ and the most enriched in disease associations^5^. Within the MHC, the classical human leukocyte antigen (HLA) genes encode the cell-surface molecules that present antigens to T cells. Identifying the HLA alleles an individual carries, known as HLA typing, is routine in immunogenetics research^6,7^ and donor-recipient matching for organ and stem cell transplantation^8–11^. Next-generation sequencing (NGS) has been rapidly adopted for clinical HLA typing^12^, but workflows vary widely in the amount of each gene sequenced and the number of MHC genes characterized.

Standard short-read variant discovery workflows, which align short reads to a linear reference, are less accurate in the MHC than other genomic regions due to the MHC’s extreme polymorphism, inter-gene sequence homology, and large structural variants^13,14^. These difficulties have prompted both specialized read-mapping tools^15^ and dedicated HLA typing algorithms^16^ that align reads directly to known HLA allele sequences^17–21^, employ graph-based alignment strategies^22–25^, or use local de novo assembly^26–29^. While HLA typing from short-read sequencing is accurate at the resolution most clinical applications require, ambiguous allele assignments remain common because short reads often cannot span the gaps between consecutive heterozygous positions, leaving multiple genotypes consistent with the data^30,31^. HLA allele ambiguities require additional testing to resolve^32^, and those that remain carry into donor selection, where genotypes are imputed from population haplotype frequencies^33,34^ and matches are predicted probabilistically rather than confirmed^35^.

Resolving the sequence of full HLA genes reveals noncoding variation that coding-only methods miss, and whether this variation matters clinically remains an open question. HLA alleles are defined by four colon-separated fields that specify increasing levels of resolution, from the amino acid sequence (fields 1–2) to synonymous coding variation (field 3) and noncoding variation (field 4), with more than 43,000 alleles cataloged in the IPD-IMGT/HLA database^36–38^. Short-read methods restricted to the antigen recognition site or coding exons cannot resolve alleles beyond three fields, and while those that sequence full-length amplicons can reach four fields, phase must be inferred across fragment gaps. Building on improvements in assembly, reference mapping, structural variant detection, and phasing^39–44^, long reads resolve phase across full genes and permit four-field, ultrahigh-resolution HLA typing^45–48^ while improving accuracy at lower fields of resolution. In retrospective studies of transplant cohorts, typing with PacBio sequencing reclassified the matching status of a substantial fraction of donor-recipient pairs previously called matched^49,50^. The clinical consequences of reclassification remain unresolved^49–52^, though pairs reclassified as mismatched have shown increased risk of grade 2-4 acute graft-versus-host disease^50^. Pairs mismatched only in noncoding regions are rare and have shown no outcome association where tested^49,50^. Noncoding variation nonetheless affects HLA gene regulation and splicing^53–55^, and may influence transplant outcomes. For example, a noncoding variant in the HLA-DPB1 3’ untranslated region has been shown to modify the risk of acute graft-versus-host disease^56^. Evaluating the consequences of noncoding HLA variation therefore requires typing methods that capture the entire gene.

Classical HLA typing characterizes only a small number of MHC genes, and the genes it omits carry clinically important variation. Even donor-recipient pairs matched at high resolution across the classical HLA loci are often mismatched elsewhere in the MHC^57,58^, including at Class III^59^, and this variation may influence transplant outcomes^58,60^. MHC Class III is among the most gene-dense regions of the human genome^7^, containing complement factors (e.g., C4A, C4B, CFB), cytokines (e.g., TNF, LTA, LTB), and numerous other genes involved in fundamental cellular processes^6^. Disease-associated variation in the MHC is not confined to the classical HLA loci. C4 copy number is associated with systemic lupus erythematosus^7,61^, and Mendelian variants in TNXB and CYP21A2 cause classical-like Ehlers-Danlos syndrome^62,63^ and congenital adrenal hyperplasia^64^. These genes lie in the RCCX module, a copy-number-variable cassette containing RP, C4, CYP21, and TNX^6,7^. High inter-gene sequence similarity in this region prevents short reads from being assigned to their locus of origin^13,65^ and collapses the region in short-read de novo assemblies^66^. C4Investigator can recover C4 copy number and variant calls^67^ but cannot phase them onto long-range haplotypes or reconstruct the full RCCX module. Resolving these genes therefore requires reads long enough to overlap sites that distinguish highly similar paralogs.

Efficient long-read HLA typing requires targeted enrichment of the MHC and typing software matched to the libraries that target enrichment produces. Most existing workflows use long-range PCR^68–71^, which offers high target specificity but can compromise accuracy through amplification bias, allele dropout, chimerism from template switching, and replication slippage in low-complexity regions^16,69,71–74^. Hybrid capture avoids gene-specific PCR primers and scales to more gene targets in a single reaction, though it takes longer, requires handling of off-target reads, and may produce uneven enrichment. While hybrid capture has been successfully applied to long-read sequencing of other immunogenetic loci^75^, existing HLA hybrid capture applications remain mostly limited to short-read platforms^66,76–78^ or to a subset of the HLA genes bundled with dozens of other non-HLA genes (e.g., [Twist Alliance Long-Read PGx Panel](https://www.pacb.com/wp-content/uploads/2023-03_PGx_Poster_Hybrid-Capture_FINAL.pdf)). On the analysis side, specialized long-read HLA typers are accumulating, but most open-source tools were designed for standard whole-genome sequencing (WGS) libraries, and in clinical settings, HLA typing is rarely performed from WGS. Amplicon and hybrid capture libraries differ from WGS in read length, coverage depth, and coverage uniformity, so a typer built for WGS input is not necessarily well matched to the data these protocols generate.

Here, we present a high-throughput target-capture approach for multiplexed long-read HLA sequencing on the PacBio and Oxford Nanopore Technologies (ONT) platforms. The workflow features one-step enzymatic fragmentation and barcoding, followed by hybrid capture with a new biotinylated probe set (Twist Bioscience) and PacBio or ONT library preparation. This workflow is readily automatable, avoids the need for expensive mechanical shearing, and is adaptable to other challenging genomic regions. We also introduce HLA-Resolve, a lightweight HLA typing algorithm for high-coverage PacBio hybrid capture libraries that returns HLA allele calls at four-field resolution. Unlike most other long-read HLA typers, HLA-Resolve requires no preprocessing other than demultiplexing, operates directly on raw FASTQ or unmapped BAM (uBAM), includes optional built-in adapter trimming, and returns clean VCFs and mapped BAMs against a modified GRCh38 reference genome. Our end-to-end workflow enables accurate reconstruction of full-length alleles for the classical HLA genes and provides comprehensive coverage of all MHC Class III protein-coding genes. We validated our approach with 32 geographically diverse samples from the Genome in a Bottle Consortium (GIAB), Human Pangenome Reference Consortium (HPRC), and International Histocompatibility Working Group (IHWG) catalogs. These datasets allowed us to validate (i) genotyping accuracy against the Genome in a Bottle benchmark, (ii) sequence identity against the phased HPRC assemblies, and (iii) HLA allele concordance against the IHWG and HPRC benchmarks. Our results demonstrate robust HLA typing performance across diverse global populations, and the resulting dataset provides a valuable resource for future method development and benchmarking.

## Results

### Hybrid capture sequencing consistently captures 85 protein-coding genes in the MHC

We developed an HLA hybrid capture assay for multiplexed long-read sequencing and applied it to genomic DNA from 33 geographically diverse human cell lines, comprising GIAB and HPRC reference samples from the Coriell Institute, as well as IHWG reference cell lines (Fig. 1, Supplementary Table 1). We tagmented the samples individually, yielding an average peak fragment size of 8,234 bases (Supplementary Fig. 1a). We pooled the tagmented samples by cohort (GIAB/HPRC, IHWG) and performed hybrid capture with a probe panel developed with Twist Bioscience, combining an existing probe set targeting twelve HLA Class I and Class II genes with a new set targeting the 69 MHC Class III protein-coding genes (Twist Design IDs LRTE-97930942 and LRTE-96945019). Post-capture libraries had an average peak fragment size of 5,637 bases (Supplementary Fig. 1b). We combined the two hybrid capture pools for library preparation and sequenced them on both the PacBio Revio (SMRTbell prep kit 3.0, 25M SMRT Cell) and the ONT PromethION (R10.4.1), loading the PromethION flow cell with approximately twice the library volume, since it generally offers higher raw throughput per flow cell.

**Fig. 1:**
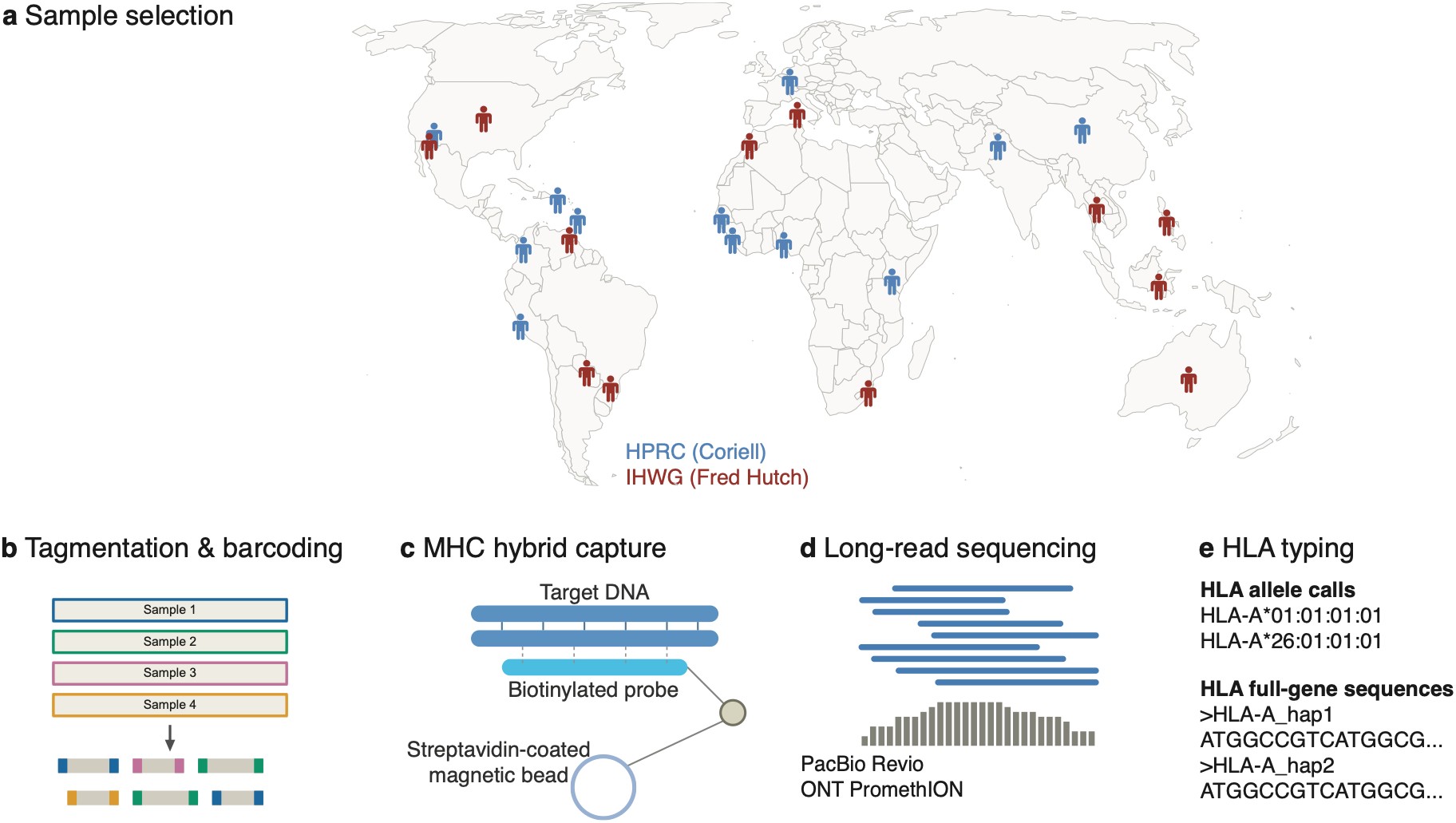
Overview of the targeted long-read sequencing workflow. **a**, Genomic DNA from 33 geographically diverse samples was obtained from two sources and enriched as two hybrid capture pools. The first comprised 17 samples from the Coriell Institute, thirteen of which are included in the first Human Pangenome Reference Consortium (HPRC) pangenome release and six of which are Genome in a Bottle (GIAB) reference samples, with HG002 and HG005 belonging to both. The second comprised 16 B-lymphoblastoid cell lines from the International Histocompatibility Working Group (IHWG) Cell and DNA Bank at the Fred Hutchinson Cancer Center. Some samples share a common ancestral origin and are represented by a single map location. **b**, Genomic DNA from each sample was tagmented and barcoded for multiplexed sequencing. **c**, Barcoded libraries were pooled by cohort for hybrid capture using a new probe set from Twist Bioscience targeting protein-coding genes in the HLA region. **d**, Post-capture libraries from both cohorts were sequenced on PacBio Revio and Oxford Nanopore PromethION platforms. **e**, HLA typing was performed with HLA-Resolve.

Following sequencing and demultiplexing, the PromethION run yielded 68,573,197 reads (163.5 Gb) and the Revio run yielded 11,337,707 reads (50.8 Gb). One sample from the HPRC cohort, HG01891, did not tagment and failed to produce sufficient sequencing reads (PacBio: 476, ONT: 1,106) and was therefore excluded from downstream analyses. Across all samples and platforms, greater than 99.7% of the raw reads mapped to the human reference genome. The number of unique (de-duplicated) mapped reads ranged from 21,397 - 448,211 for PacBio, and 183,480 - 1,475,328 for ONT (Fig. 2a). Mean and median read lengths for target-overlapped reads were longer for PacBio (mean=4,093, median=3,910, N50=4,331) than ONT (mean=2,804, median=2,693, N50=3,306) (Fig. 2b, Supplementary Fig. 2). On-target mean coverage depth was high on both platforms (PacBio 148x, ONT 286x) (Fig. 2c), with fold enrichment, defined as the ratio of mean on-target to off-target depth, ranging from 505 to 921x (Fig. 2d). Target specificity was consistent across hybrid capture pools and across samples within a hybrid capture pool (Supplementary Fig. 3, Supplementary Tables 2 and 3), and variation in coverage depth among samples within a cohort was primarily driven by sequencing effort (i.e., reads per sample) rather than by variation in target specificity (Fig. 2a, Supplementary Fig. 3). Differences in coverage depth between cohorts are detailed in Supplementary Note 1. Within each cohort, raw read counts per sample were highly correlated between PacBio and ONT (HPRC: Pearson r = 0.89; IHWG: Pearson r = 0.96), as was mean coverage depth across captured protein-coding genes (Fig. 2e,f, Fig. 3), indicating consistent performance across platforms at both the sample and gene level.

**Fig. 2:**
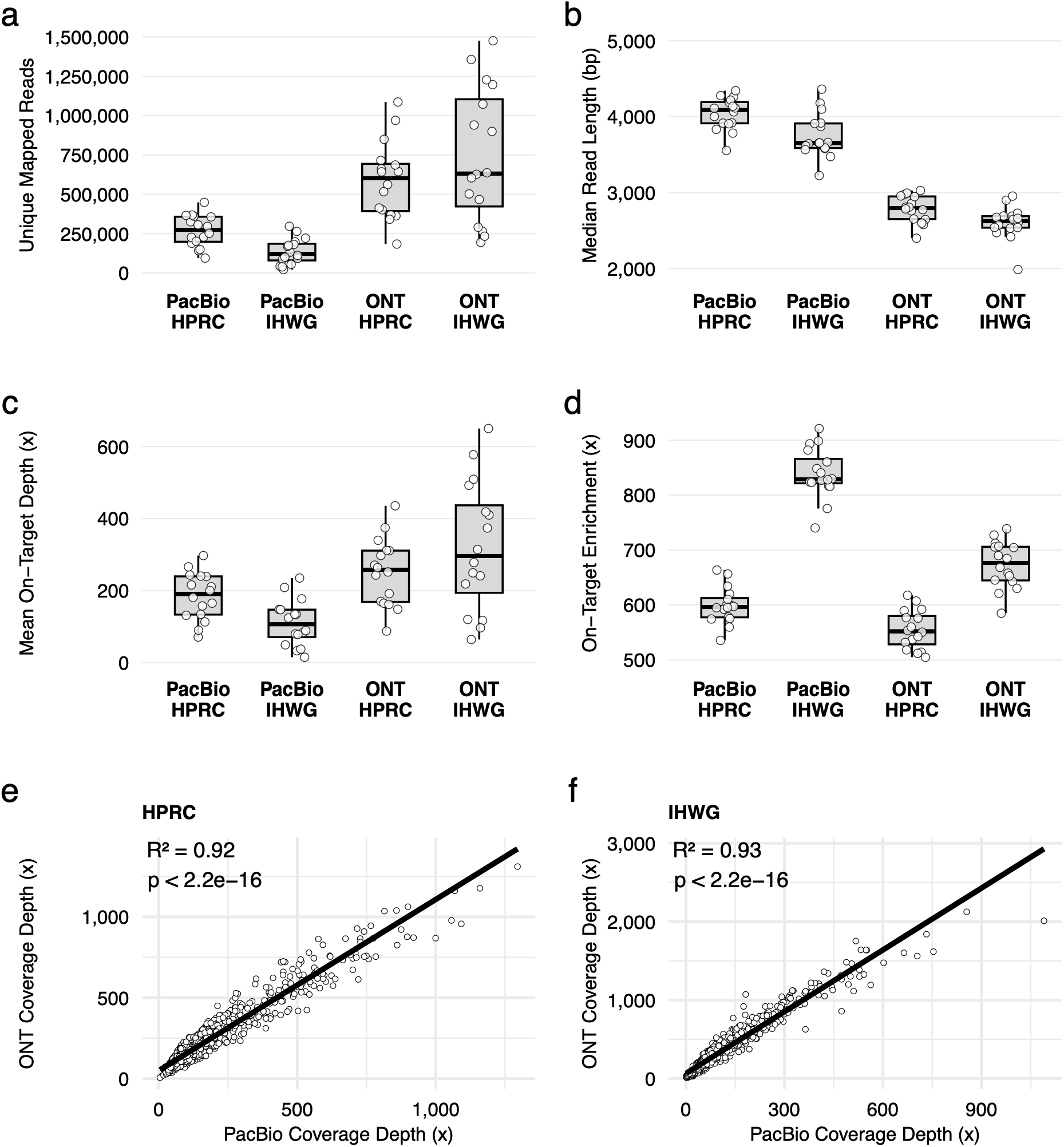
Summary of targeted long-read sequencing performance. **a,** Number of mapped sequencing reads per sample, with PCR duplicates removed. **b,** Median read length of target-overlapped primary alignments. **c,** Mean on-target coverage depth (x). **d,** On-target enrichment, defined as the ratio of mean on-target to off-target coverage depth. For panels a–d, results are stratified by sequencing platform (PacBio or ONT) and hybrid capture cohort (HPRC or IHWG), and points represent individual samples. **e–f**, Comparison of ONT versus PacBio coverage depth across 83 consistently captured protein-coding genes for matched samples from the HPRC (e) and IHWG (f) cohorts. Each point represents a gene-sample pair. The solid line shows the linear regression fit, with R² and p-values annotated.

**Fig. 3:**
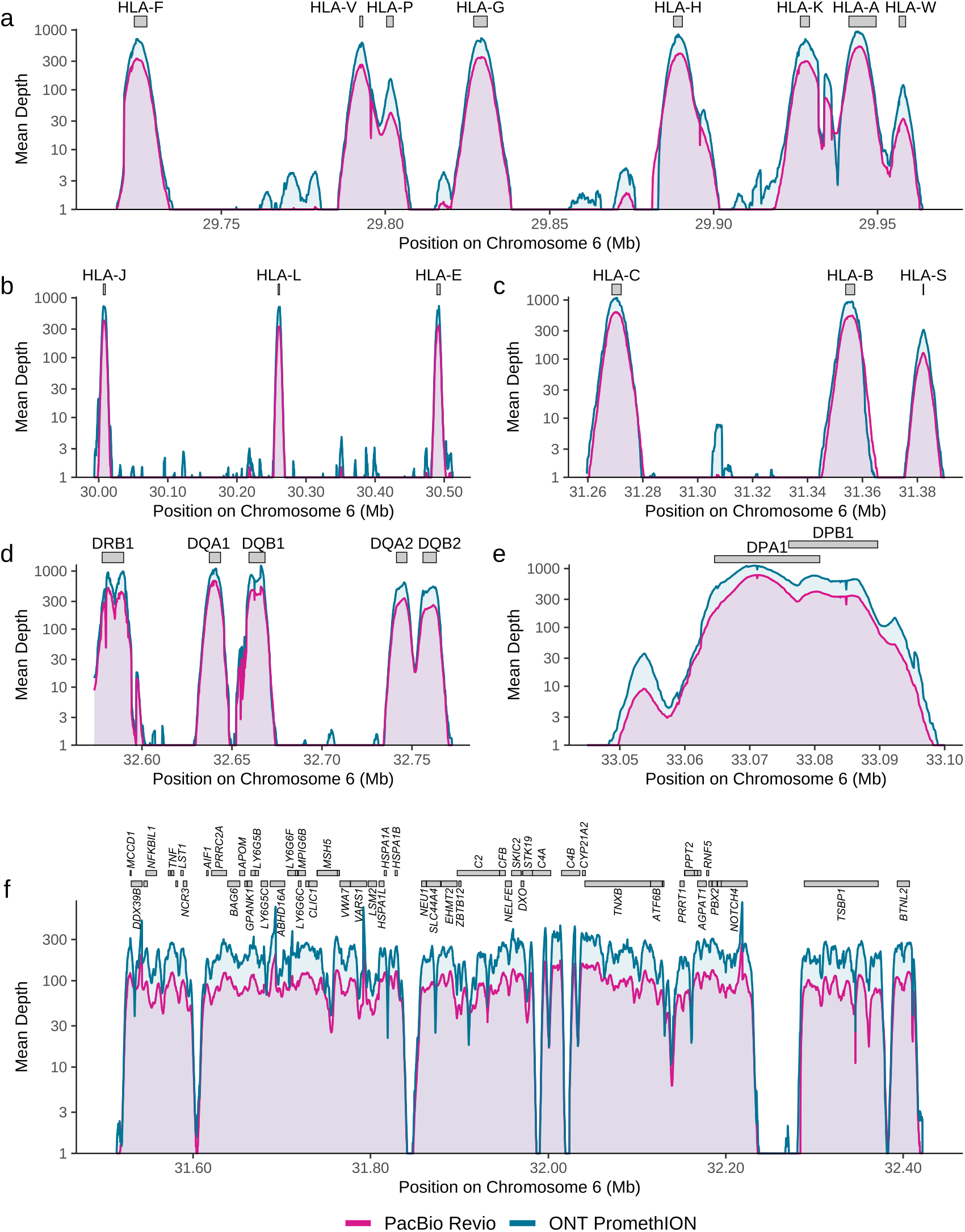
Average coverage depth across selected HLA regions. Mean coverage depth (log scale) across the MHC region is shown for PacBio Revio (magenta) and ONT PromethION (teal), using per-base coverage averaged across samples and aggregated into non-overlapping 100 bp windows. **a-c,** The HLA Class I region, including HLA-A, HLA-B, HLA-C, and selected non-classical HLA genes and pseudogenes. **d-e,** The HLA Class II region. **f,** The entire Class III region. Coverage gaps reflect intergenic regions not targeted by the hybrid capture probes, except for the two small gaps flanking 32.00 Mb, corresponding to the C4A and C4B intron 9 HERV insertion. In f, plus-strand genes are on the upper row and minus-strand genes on the lower row, with only protein-coding genes shown. Windows with mean depth below 1x are plotted at 1x because the y-axis is on a log scale.

We consistently captured all 81 targeted protein-coding genes, including the 69 Class III genes, along with four untargeted protein-coding genes, HLA-E, HLA-F, HLA-G, and HLA-DQB2, providing coverage for 85 protein-coding genes in total (Fig. 3, Supplementary Figs. 4 and 5). HLA-DRB3 and HLA-DRB4 lie outside the GRCh38 primary assembly, so we confirmed their capture by competitive mapping against a multi-allele DRB reference, which recovered reads in every sample whose HLA-DRB1 genotype predicts the corresponding paralog. We also captured numerous pseudogenes, lncRNAs, snoRNAs, and miRNAs annotated in the human reference genome, including HLA-H, HLA-J, and HLA-K. See Supplementary Tables 4–11 for per-sample mean coverage depth and proportion of bases covered by 20 or more reads, reported by gene and exon for both PacBio and ONT.

Although the read lengths generated from the hybrid capture protocol are shorter than those of standard PacBio and ONT whole-genome sequencing, the reads were sufficiently long to distinguish between highly paralogous genes, such as protein-coding gene CYP21A2 and its pseudogene paralog CYP21A1P (Fig. 4a). Our reads were also able to distinguish C4A from C4B, and differentiate between long and short isoforms with or without the long intron 9 human endogenous retrovirus (HERV) insertion (Fig. 4a). Across the HLA region, we were able to detect deletions up to 6,367 bp and insertions up to 3,655 bp (Fig. 4b, Supplementary Table 12) using pbsv with the PacBio reads. To confirm read-level support for detected structural variants, we manually inspected all variants >500 bp in Integrative Genomics Viewer^79^ (IGV) and removed those lacking supporting reads. The protein-coding genes harboring the most structural variants (insertions/deletions >= 50 bp) were the HLA Class II genes HLA-DRB1, HLA-DRB5, HLA-DQA1, and HLA-DQB1, and the Class III genes TSBP1, C4A, C4B, and C2, consistent with the known structural complexity of the Class II loci and the RCCX module of Class III (Fig. 4c). Structural variants were distributed relatively evenly across samples, ranging from 6 to 49 per sample (median = 33), with no individual samples disproportionately contributing to the total call set (Fig. 4d).

**Fig. 4:**
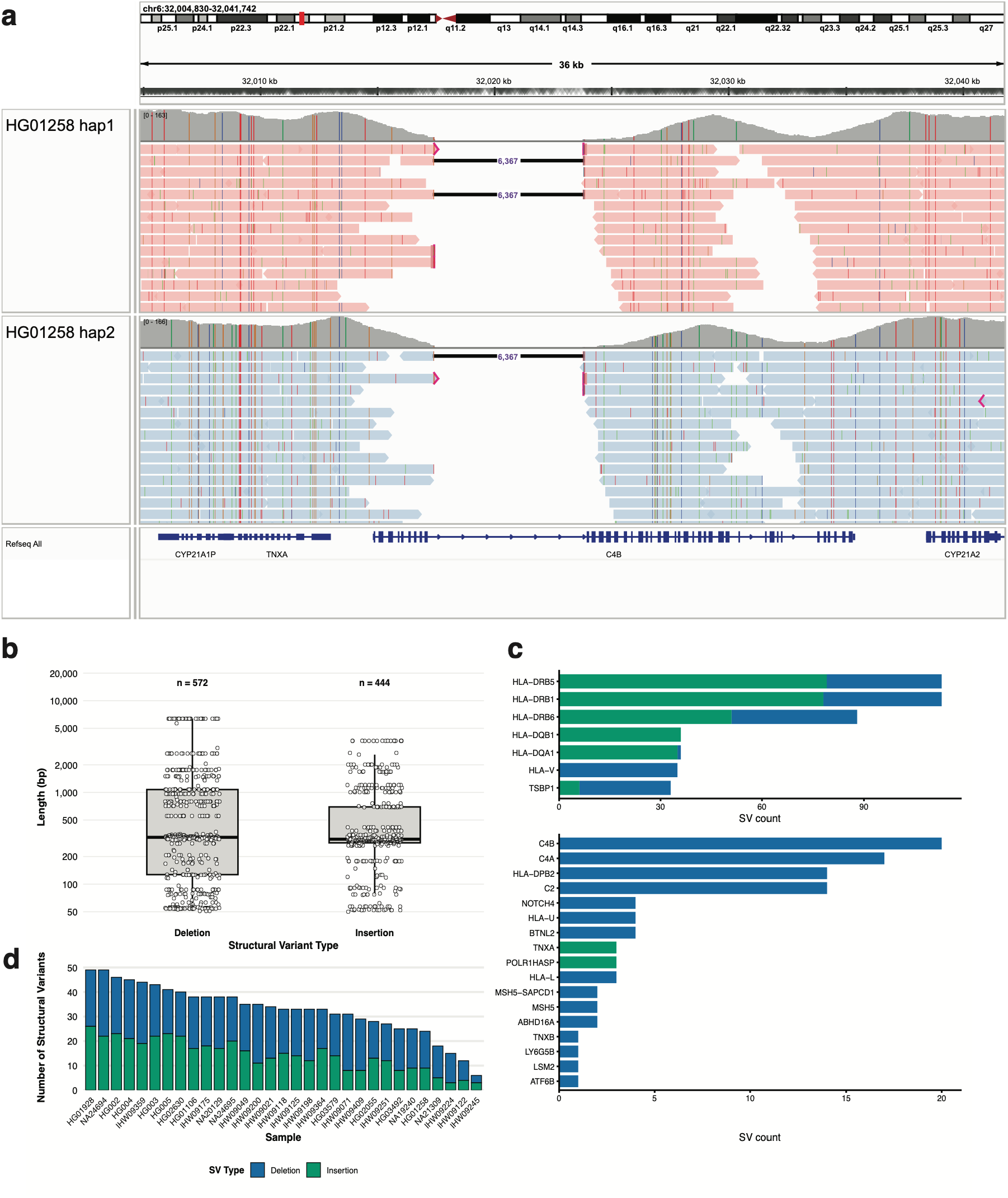
Characterization of CYP21A2 read mapping and MHC-wide structural variation. **a,** An Integrative Genomics Viewer (IGV) plot of sample HG01258 showing primary alignments to CYP21A2 and CYP21A1P, with reads separated by haplotype. Also shown is a large 6,367 bp deletion in C4B, representing the short C4B isoform lacking the intron 9 HERV insertion. **b**, Distribution of structural variant (SV) lengths by type. Each point represents one SV in an individual sample. The y-axis is log10-scaled. Total SV counts across all samples are shown above each column. **c**, Number of SVs overlapping annotated genes, stratified by SV type. The plot is split into two panels with different x-axis scales: x >= 30 (top) and x < 30 (bottom). **d**, Total SV count per sample, stratified by SV type. Samples are ordered by decreasing total SV count. For **b-d**, only SVs greater than 49 base pairs in length were included. Sample IHW09117 was excluded (Supplementary Note 2), so panels b-d show 31 samples.

### Clinical-level small variant accuracy achieved with HiFi reads

To evaluate small-variant genotyping accuracy, we called variants with DeepVariant and Clair3 and compared the results to the Genome in a Bottle (GIAB) high-confidence benchmarks (v4.2.1). For the ONT reads, we evaluated both DeepVariant and Clair3, and Clair3 performed better. We therefore used Clair3 for all downstream ONT analyses. We assessed genotype concordance for samples HG002–HG007 using hap.py (https://github.com/Illumina/hap.py) following the GA4GH/GIAB best practices^80^. We restricted analyses to GIAB’s confident regions and loci targeted by our hybrid capture probes, resulting in 2,113 (HG003) to 3,146 (HG007) truth SNVs and 177 (HG002) to 250 (HG007) truth indels. We observed strong genotype concordance from both platforms, with PacBio + DeepVariant achieving clinical accuracy (mean SNV F1 = 99.8%, mean indel F1 = 98.4%) and outperforming ONT + Clair3 (SNV F1 = 98.4%, indel F1 = 94.1%; Fig. 5a, Supplementary Table 13). Discordant calls for both platforms were concentrated in C4A, C4B, and the HLA-DQ genes, with HLA-DQ accounting for the majority of false positives and false negative genotypes. The distribution of SNVs in the GIAB benchmark is not uniform across our targeted regions, as SNVs are not evenly distributed among HLA genes. For example, 74.5% (2,467/3,312) of the HG002 high-confidence GIAB SNVs fall within the captured gene bodies of HLA-A, HLA-B, HLA-C, HLA-DQA1, HLA-DQB1, and TSBP1 (Supplementary Fig. 6). Furthermore, HLA-DRB1 is not included in GIAB’s confident regions, so we were not able to evaluate genotype concordance within this gene.

**Fig. 5:**
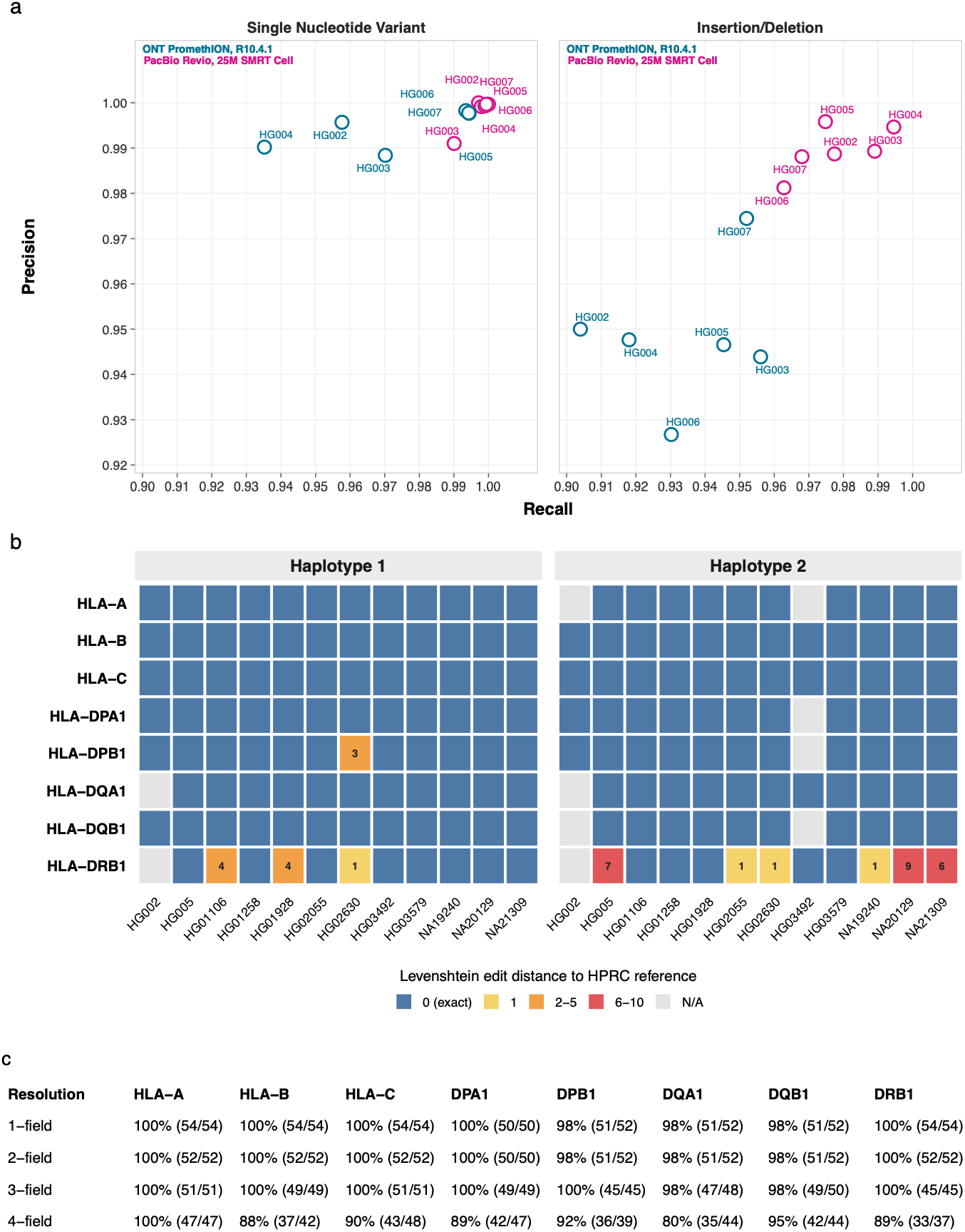
MHC genotyping and HLA allele typing accuracy: **a,** Single-nucleotide variant (left) and indel (right) genotype concordance with the Genome in a Bottle (GIAB) benchmark for HG002–HG007, colored by sequencing platform. Variants were called with DeepVariant v1.6.1 for PacBio reads and Clair3 v2.0.0 for ONT reads. Precision and recall are shown for each sample, computed across captured protein-coding genes within the HLA region. **b**, Heat map of Levenshtein edit distance between each reconstructed HLA allele sequence and its corresponding Human Pangenome Reference Consortium (HPRC) assembly sequence, for the PacBio-sequenced samples with an available assembly. Distances were computed with edlib in infix alignment mode across the gene sequence, including introns and excluding 5’ and 3’ UTR. Reference sequences were extracted from the HPRC Release 2 assemblies and CAT annotations, except HG002 and HG03492, which have no Release 2 assembly, for which we used the Ensembl-annotated Release 1 assembly. Cells are colored by binned edit distance, with exact matches in blue and increasing distances on a warm scale. The edit distance value is annotated in text for cells with non-zero values. Labeling of haplotypes 1 and 2 is arbitrary and does not denote a consistent parental phase. Gray cells indicate gene and haplotype combinations absent from the reference annotations. **c,** HLA allele concordance of PacBio HLA-Resolve calls with the combined 12 HPRC and 15 IHWG reference typings, stratified by gene and resolution level.

To determine the minimum sequencing depth required for accurate genotyping, we iteratively downsampled the HG002 PacBio reads and recalculated coverage depth and genotype concordance with the GIAB benchmark. Because coverage depth varied across HLA classes, we downsampled and evaluated concordance separately for each HLA class. To account for sampling variance, we generated ten independent downsampling replicates at each coverage level, and computed mean recall, precision, and F1 score (Supplementary Figs. 7 and 8, Supplementary Table 14). As coverage depth decreased, recall declined sharply. In contrast, precision remained consistently high, reflecting accurate genotyping at the subset of variant sites that were still detected. The depth required for 98% SNV recall differed by HLA class. Recall at Class I and II varied substantially among downsampling replicates, exceeding 98% in all ten replicates only at 60x depth for Class I and at 153x for Class II, though individual Class II replicates still occasionally fell below 98% at higher depths (Supplementary Fig. 8). Class III maintained SNV recall above 98% for all replicates at approximately 12x.

### High-accuracy gene sequence reconstruction enables high-resolution HLA typing with HLA-Resolve

We developed HLA-Resolve, a Python command-line tool that performs four-field HLA allele calling directly from raw PacBio reads (FASTQ/uBAM). HLA-Resolve follows a standard variant-discovery workflow, including optional adapter trimming, optional PCR duplicate removal, reference-genome alignment, variant calling, structural variant calling, tandem repeat genotyping, and genotype phasing. The phased genotypes are then applied to the human reference genome to reconstruct coding and full-gene phased sequences, which are then queried against the IPD-IMGT/HLA database to identify the best-matching allele by sequence similarity. In addition to the reconstructed nucleotide sequences and allele assignments, HLA-Resolve outputs haplotagged BAMs, phased VCFs, and coverage depth metrics.

We ran HLA-Resolve v0.9.9 on all HPRC and IHWG PacBio samples, evaluating sequence reconstruction accuracy against the HPRC phased assemblies and HLA allele concordance against the HPRC and IHWG reference typings. We excluded one IHWG sample (IHW09117) due to contamination signatures (see Supplementary Note 2). Comparison of our PacBio full-gene sequences to the HPRC assemblies showed high accuracy (Fig. 5b). Of 182 reconstructed allele sequences, 172 (94.5%) had zero edit distance to the corresponding assembly, including every allele sequence at HLA-A, -B, -C, -DPA1, -DQA1, and -DQB1 (Fig. 5b, Supplementary Table 15). Only ten alleles differed at all, nine at HLA-DRB1 and one at HLA-DPB1, and the largest difference was nine bases. Of 496 reconstructed coding sequences, 478 (96.4%) exactly matched an allele in the IPD-IMGT/HLA database, and a further 17 differed only at ambiguous bases introduced during DR and DQ re-consensus (see Methods), leaving 495 (99.8%) identical at every called base.

Across 15 IHWG samples, HLA allele concordance was 98.6% (219/222) at two-field, 99.0% (196/198) at three-field, and 83.3% (135/162) at four-field resolution. For the 16 HPRC samples, concordance against the HPRC reference typings established by Lai et al.^78^ was higher, reaching 100% at two-field (192/192) and three-field (190/190) resolution, and 96.8% (180/186) at four-field resolution. Pooled across both cohorts, HLA-Resolve was 99.3% concordant at two-field (411/414), 99.5% at three-field (386/388), and 90.5% at four-field resolution (315/348) (Fig. 5c). Among the PacBio samples, HLA-Resolve v0.9.9 outperformed FuFiHLA^48^, StarPhase^46^ and SpecImmune^47^ at all fields of resolution (Fig. 6, Supplementary Table 16). FuFiHLA was only compared across the six genes it types, since it does not type HLA-DPA1 or HLA-DPB1. HLA-Resolve is currently PacBio-specific, so we benchmarked FuFiHLA, StarPhase, and SpecImmune on ONT reads, where all three performed comparably to their PacBio results. When pooled across both cohorts, ONT concordance at three- and four-field resolutions was about 3 percentage points lower than PacBio for StarPhase, 2 percentage points lower for SpecImmune, and within 1 percentage point for FuFiHLA (Fig. 6). We also evaluated HLA*LA^24^ and observed strong performance and compatibility with our reads, but did not quantify HLA allele concordance, as HLA*LA returns calls at G group resolution.

**Fig. 6:**
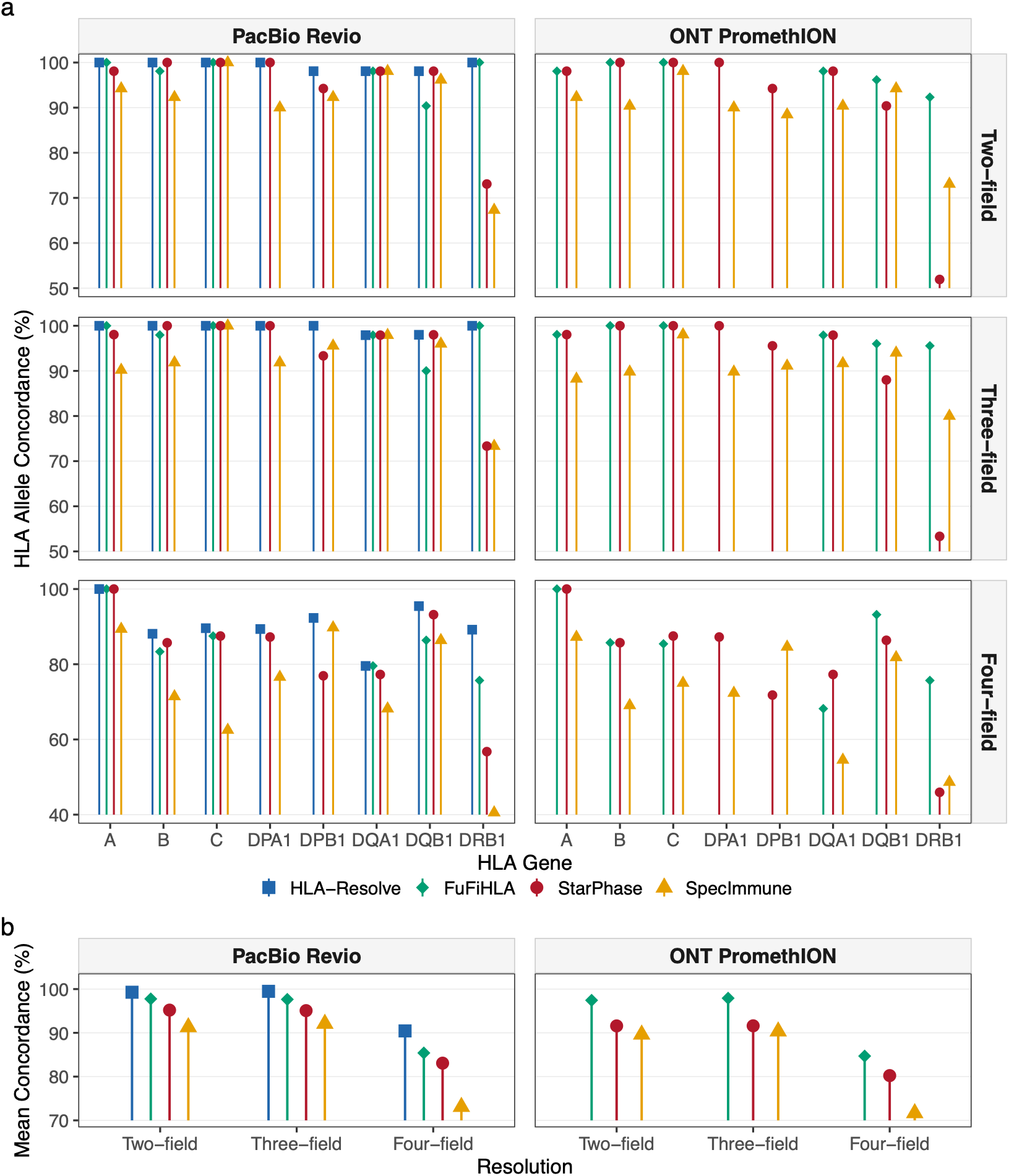
HLA allele concordance with the HPRC and IHWG reference typings. **a,** HLA allele concordance (%) with HPRC and IHWG reference typings for HLA-Resolve (blue squares), FuFiHLA (green diamonds), StarPhase (red circles), and SpecImmune (orange triangles), shown for each HLA gene and stratified by typing resolution (two-, three-, or four-field) and sequencing platform (PacBio Revio, ONT PromethION). Each point represents the concordance for a given gene across all 27 samples (15 IHWG, 12 HPRC). HLA-Resolve results are shown for PacBio Revio only. For FuFiHLA, concordance is omitted for DPA1 and DPB1, which it does not type, and the mean is taken over the six genes it types. **b**, Unweighted mean of the per-gene concordances in a, summarized by resolution and platform. Each gene contributes equally regardless of the number of testable alleles.

We benchmarked runtime and memory usage for our hybrid capture data on a dual-socket AMD EPYC 7663 node running AlmaLinux 9.7, processing each sample with 6 CPU threads and allocating 25GB of RAM. The 31 processed PacBio samples had a median of 316,846 raw reads each (range 42,221 to 888,440). HLA-Resolve had a mean wall time of 9.3 minutes (range 4.3 to 14.7 minutes) and a peak resident memory usage of 15.1 GB.

### HLA typing accuracy extends to whole-genome sequencing libraries

To test whether HLA-Resolve generalizes beyond hybrid capture input, we applied it to 146 public PacBio WGS libraries from 130 samples sequenced on the Revio or Sequel II, with a mean coverage depth of 33.1x across the eight classical HLA genes (median 33.9x, range 19.6-46.8x). We typed each library separately and treated two libraries of the same sample as independent replicates. Of the 130 samples, 118 were not included in our hybrid capture cohorts. For the 43 HPRC release 1 samples (52 libraries), HLA allele concordance with the reference typings of Lai et al.^78^ was 99.8% (829/831) at two-field, 99.5% (816/820) at three-field, and 93.0% (745/801) at four-field resolution (Supplementary Table 17). For the remaining 87 samples (94 libraries), concordance with the 1000 Genomes Project HLA panel was 99.0% (903/912) at two-field resolution, which is the panel’s limit. Across all 146 libraries, the call rate was 100% (2,336/2,336 loci), and two-field concordance was 99.4% (5,196/5,229). HLA-Resolve therefore accurately types HLA genes from whole-genome data, with no change to the workflow.

### Full-gene sequence reconstruction requires greater coverage depth than allele typing

To assess how sequencing coverage affects HLA typing performance, we downsampled aligned reads for each sample to a series of target depths and retyped the samples at each depth. We split the aligned reads by HLA gene and downsampled each gene separately to control for variation in coverage depth among genes within a sample, and we performed ten replicates at each depth level to control for variation from random read subsampling. Here, depth refers to the length-weighted mean coverage across each gene’s coding sequence. We report the realized depth in each downsampled alignment rather than the target depth because random subsampling does not exactly produce its target depth. HLA allele concordance was computed for each gene at each depth, across all alleles from the 21 samples that reached every target depth, then averaged over the ten replicates. At two- and three-field resolution, every gene came within one percentage point of its full-depth concordance (the same samples typed with all reads) between 12x and 25x (Supplementary Fig. 9). Four-field typing converged more slowly. HLA-C came within one percentage point of full-depth concordance at 30x, while HLA-B and HLA-DRB1 remained 1.6 and 2.1 points below full-depth concordance at 200x.

We also measured sequence reconstruction accuracy at each depth as the edit distance between the downsampled reconstruction of each gene and its full-depth reconstruction in the same sample. Across genes, the median coding sequence reconstruction became identical to full depth at 10-15x, and 90% of reconstructions across samples and replicates matched full depth exactly at 13-30x (Supplementary Figs. 10a,b). Whole-gene reconstruction, including noncoding regions, converged far more slowly. The median reconstruction was within one edit of the full-depth reconstruction by 15x for HLA-C, 20x for HLA-DQA1 and -DQB1, 23x for HLA-DPA1 and 35x for HLA-DPB1, but not until 124x for HLA-A. HLA-B and HLA-DRB1 retained median distances of 24 and 2 even at 200x. For four genes, 90% of reconstructions were identical to the full-depth reconstruction within the tested range: 35x for HLA-C, 60x for HLA-DQB1, 64x for HLA-DPA1, and 107x for HLA-DPB1. HLA-A, -B, -DQA1, and -DRB1 did not reach this threshold. HLA-A and HLA-B performed worse because their annotated transcripts are only 40% and 24% of their reconstructed gene records, and the sequence outside the transcript is covered at 5-fold lower depth than the coding exons for HLA-B and 2-fold lower for HLA-A. Therefore, our CDS-normalized downsampling left these regions with less coverage than reported. For example, at the HLA-B 200x level, the sequence outside the transcript had a mean depth of only 37x.

To separate reconstruction accuracy from coverage of the sequence outside the annotated transcript, we compared an additional interval, defined as the sequence between the start and stop codons with introns retained. For these comparisons, the median reconstruction was identical to full depth from 10x for HLA-A, -B, and -C and 13x for HLA-DPA1, but required 20x for HLA-DQA1, 22x for HLA-DQB1, and 35x for HLA-DPB1. HLA-DRB1 is the exception. Although its coding sequence was reconstructed exactly in 97.2% of records at 200x, its start-to-stop interval retained a median edit distance of 2 and matched the full-depth reconstruction exactly in only 35.3% of records, reflecting the difficulty of reconstructing introns 1, 2, and 5.

Because sequence reconstructions can also be truncated at low depth when phasing is interrupted, we measured the proportion of coding sequences shorter than the full-depth reconstruction. Truncation was at or near zero at every depth for HLA-A, -B, -C, and -DPA1, and fell to 1% or less by 18x for HLA-DQB1, 30x for HLA-DQA1, and 89x for HLA-DPB1. HLA-DRB1 fell from 31% at 10x to 1.2% at 200x (Supplementary Fig. 10c).

### The targeted sequencing workflow is reproducible across independent hybrid captures and sequencing runs

We participated in the 19^th^ International HLA and Immunogenetics Workshop (IHIWS) project “NGS of full-length HLA genes of reference cell lines,” in which independent laboratories performed HLA typing on blinded DNA panels from the IHWG B-lymphoblastoid cell line collection to create consensus full-field reference typings. We captured and sequenced 96 samples across eight hybrid capture pools and two PacBio Revio runs, using the same protocol as the main cohort. We included two samples from our main cohort (HG002, IHW09224) as inter-run replicates. The remaining 94 samples were from two quality-control and four unknown workshop panels. Consensus typings for these panels have not yet been released, so we used this set to assess reproducibility of the targeted sequencing assay only.

One of the eight hybrid capture pools yielded only 5.7 ng of post-capture library compared with 148-215 ng for the three other pools in the same run. We sequenced this pool but excluded it from downstream analyses as it only produced a median of 573 unique mapped reads per sample, too few for HLA typing (Supplementary Table 18). Of the 84 samples from the other seven pools, 81 (96%) had sufficient reads for HLA typing, and HLA-Resolve called 1,292 of the 1,296 (99.7%) possible alleles (Supplementary Tables 19 and 20). All allele calls were resolved to the full resolution available in the IPD-IMGT/HLA database (Supplementary Tables 21 and 22). The four uncalled alleles were HLA-DQA1 in two samples that fell below HLA-Resolve’s requirement of 8x mean coverage depth over the antigen recognition site. Of the 646 called loci typed across the 81 samples, 631 had a mean CDS depth of at least 20x, which we showed was sufficient for high-accuracy typing at two- and three-field resolution in our downsampling analysis. The two re-run samples from our main cohort gave identical allele calls in both runs at all eight classical HLA loci (Supplementary Table 23).

Target specificity varied between hybrid capture pools (Supplementary Fig. 11). Two pools from the first sequencing run, captured as a separate batch, had twice the post-capture yield and on-target rates of 1.5% - 1.7% compared with 20-31% of the other pools, consistent with off-target carryover during hybridization or washing. However, median ARS depth at the eight HLA genes in these pools was still 43-232x, and HLA-Resolve typed every locus in 22 of the 24 samples. In the second sequencing run, Class II showed reduced depth, especially at HLA-DQA1, with Class I unaffected. Wash stringency is the likely cause, as HLA-DQA1 has the lowest GC content of the eight classical HLA genes, and therefore forms the least stable probe-target duplexes. We are evaluating Twist Bioscience’s updated recommendations for wash temperature and step-down denaturation. The reduced HLA-DQA1 depth in the second run accounts for the two uncalled loci described above. All other loci were called in both runs.

## Discussion

We present an end-to-end long-read HLA typing workflow that pairs single-step tagmentation and barcoding with a hybrid capture panel that enriches the classical and non-classical HLA genes and all Class III protein-coding genes, and introduce HLA-Resolve, an HLA typing tool that outperforms existing open-source long-read typers on the HiFi reads this workflow produces. Our one-step tagmentation and barcoding method consistently generates fragments of the size and yield required for hybrid capture and multiplexed, platform-agnostic long-read sequencing. We also demonstrate consistent target enrichment across all 32 samples in two independent hybrid capture experiments. Our protocol is readily automated on standard liquid handlers and eliminates the manual mechanical shearing step (e.g., Diagenode Megaruptor or Covaris g-TUBE), reducing costs, improving reproducibility, and enabling sample tracking that minimizes human error. Although our captured fragments were shorter than typical PacBio and ONT WGS libraries, they were sufficient for high-accuracy genotyping and HLA allele assignment. We benchmarked three high-resolution open-source long-read typers, FuFiHLA, SpecImmune, and StarPhase, on both PacBio and ONT reads, all of which showed strong concordance on both platforms. For PacBio reads, our tool HLA-Resolve outperformed all three and provides a fully automated bioinformatic workflow compatible with any PacBio library with high coverage across the HLA genes. HLA-Resolve successfully reconstructed phased full-gene sequences with a standard variant-discovery workflow and accurately matched them to the IPD-IMGT/HLA database using a biologically interpretable edit distance metric. The high-coverage HiFi reads produced by our sequencing protocol support a conventional linear reference-based implementation that is lightweight, fast, and intuitive, avoiding the need for de novo assembly or graph-based alignment. By reconstructing allele sequences before database matching, the inferred gene sequences for HLA-A, -B, -C, -DPA1, and -DPB1 are not influenced by the composition of the IPD-IMGT/HLA reference set. Three Class II genes, HLA-DQA1, HLA-DQB1, and HLA-DRB1, do receive a second consensus sequence built by re-mapping the reads to the best-matching database allele. However, this step refines only the reconstructed sequence and the fourth field, since the search is confined to fourth-field options within the assigned three-field allele group. While we developed and tested the protocol on the HLA region, the underlying targeted sequencing protocol is generalizable to any genomic region by modifying the hybrid capture probe panel.

In addition to HLA typing, our method provides comprehensive coverage of the Class III region, which contains many clinically important genes that are poorly served by existing approaches. The most structurally complex of these is the RCCX module, a multi-gene cassette whose copy-number and structural variation underlies several Mendelian and complex diseases. For example, CYP21A1P-CYP21A2 fusion deletion mutations cause congenital adrenal hyperplasia^64^, low C4 copy-number is associated with systemic lupus erythematosus^7,61^, and TNXB deficiency causes classical-like Ehlers-Danlos syndrome^62,63^. These variants are difficult to detect with short-read sequencing due to the high sequence identity between paralogous genes and pseudogenes. The long reads generated by our approach can be reliably assigned to CYP21A2 or the CYP21A1P pseudogene and are compatible with specialized tools such as Paraphase^65,81^ for resolving pathogenic variants. Reference alignment at C4A and C4B remains challenging, but our long reads anchor to the sites that distinguish the two genes in GRCh38 and may be well suited for local de novo assembly. Together, the sequencing strategy and available analytical tools provide a promising route to studying RCCX-associated disease in parallel with HLA typing.

While our protocol successfully captures all targeted genes, we are working to improve two aspects of the capture design: the ratio between the two probe sets, and the scope of the panel. Across the main cohort, the mean coverage of HLA Class I and II genes was three- to fourfold higher than that of Class III genes, reflecting the 1:1 ratio at which we pooled the two biotinylated probe sets. We will adjust this ratio in future iterations of the assay. We are also developing a second version of our probe panel with expanded scope, adding further MHC, killer cell immunoglobulin-like receptor (KIR), and leukocyte immunoglobulin-like receptor (LILR) gene targets.

Our reference-based approach to HLA typing has three main limitations. The first is reconciling overlapping variant calls from multiple callers (e.g., DeepVariant, pbsv), which may genotype the same variants but annotate them differently. The second concerns genes with high structural variation, particularly HLA-DRB1, whose first intron is long and structurally variable and frequently carries kilobase-scale insertions relative to the reference genome. GRCh38 contains one of the shortest HLA-DRB1 alleles, DRB1*15:01:01:01 (https://www.ebi.ac.uk/ipd/imgt/hla/help/genomics.html). Across the IPD-IMGT/HLA catalog, intron 1 length spans several kilobases, from 5,262 bases in DRB1*15:01:01:07 to 10,315 bases in DRB1*09:01:02:05. Insertions of this scale complicate alignment, variant calling, and phasing continuity whenever few reads fully span the inserted sequence, and we frequently observed a phasing break at the position of the insertion, most likely because the probes did not directly target these intron sequences due to high repeat content. The third arises in the DR cluster, where the Twist probes appear to capture fragments from untargeted HLA-DRB paralogs whose reads can mis-map onto HLA-DRB1 and corrupt allele calling.

HLA-Resolve addresses all three problems at successive stages of the workflow. Reads from the DR cluster are first remapped against a multi-allele HLA-DRB reference and discarded if their primary alignment falls outside HLA-DRB1, thereby removing paralog contamination before genotyping. Redundant genotypes are screened out before sequence reconstruction (see Methods). A final re-consensus step then rebuilds HLA-DQA1, HLA-DQB1, and HLA-DRB1 allele sequences from reads re-aligned to the best-guess allele, with the database search restricted to fourth-field options inside the already-assigned three-field family, so refinement is confined to the fourth field, and lower-resolution calls are never overturned. This step is designed to bridge phasing interruptions left in the HiPhase output. Together, these steps yielded HLA-DRB1 concordance of 100% at three-field and 89% (33/37) at four-field resolution. Increasing the number of reads that fully span non-reference insertions remains a priority for contiguous full-gene reconstruction, although their high repeat content^82^ may increase off-target enrichment. Local assembly is an alternative but remains unreliable for HLA genes, even with read lengths far exceeding those generated by our tagmentation-based library prep. Across the 24 HPRC v1 haplotype assemblies from the 12 HPRC samples in this study, 82 of 192 (42.7%) classical HLA gene annotations were absent from the Ensembl GFF3 annotation, possibly due to assembly gaps, a problem also seen in the high-coverage 1000 Genomes Project (1KGP) ONT assemblies from Gustafson et al.^83^. The HPRC release 2 CAT annotations recover nearly all of these, so we used Release 2 assemblies where available.

The commercial HLA typing landscape is rapidly evolving toward targeted long-read sequencing. Most current assays use either Illumina-based hybrid capture (e.g., CareDx AlloSeq Tx 17, Thermo Fisher One Lambda HybriType NGS Assay, Omixon Holotype HLA) or ONT-based amplicon sequencing (e.g., Thermo Fisher AllType Rapid Assay, Omixon NanoTYPE, GenDx NGS-Pronto, and CareDx AlloSeq Nano), but new long-read hybrid capture options are emerging, including the QIAGEN QIAseq xHYB HLA panel for PacBio or ONT sequencing and GenDx’s ONT-based NGS-Capto. Despite this progress, transparent benchmarking remains limited across the field. External proficiency testing programs administered by ASHI, CAP, and EFI assess clinical laboratory performance, but, to our knowledge, no published results allow for comparison across assays. Multi-laboratory studies have typed IHWG reference cell lines across a range of commercial platforms^84,85^, but these studies were designed to establish consensus genotypes rather than to rank methods. Laboratory identities were dissociated from performance, and concordance was scored against consensus derived from the participating laboratories themselves. No published study therefore permits a head-to-head comparison of typing solutions on the same samples. Vendors participate in the International HLA and Immunogenetics Workshop and contribute genotyping data but rarely publish detailed performance metrics. Academic HLA typing workflows face similar benchmarking concerns. Custom sequencing protocols are commonly paired with commercial analysis software such as GenDx NGSengine or Omixon HLA Explore, but these hybrid strategies are rarely validated against public benchmarking datasets. Performance is more often reported as rates of ambiguous calls or the proportion of allele assignments at different resolution levels, with reproducibility emphasized over concordance against independent truth sets. This lack of standardized benchmarking across both commercial and academic settings limits our ability to identify the major unresolved challenges in high-resolution HLA typing and to direct efforts toward the most pressing problems.

The proprietary nature and cost of commercial assays further limit their accessibility in academic research, motivating the development of open, cost-effective alternatives. Our assay captures 85 protein-coding genes at approximately $100 per sample in consumables, covering library prep, hybrid capture, and sequencing, when 33 samples are multiplexed per flow cell. Per-sample costs should continue to fall with higher multiplexing and recent chemistry improvements (SPRQ-Nx), while benchtop instruments such as the PacBio Vega also make PacBio HLA typing more accessible. Our downsampling analysis measuring the relationship between coverage depth and HLA typing accuracy provides a practical target for multiplexing. Two- and three-field concordance came within one percentage point of full-depth performance for every gene between 12x and 25x coding-sequence depth, and 90% of coding sequences matched the full-depth reconstruction exactly by 13x to 30x. Four-field typing and whole-gene reconstruction converged more slowly, and four-field concordance for HLA-B and HLA-DRB1 remained below full-depth performance even at 200x, which reflects the thinner coverage of noncoding sequence outside the annotated transcript and the difficulty of reconstructing the HLA-DRB1 introns. Laboratories that require only three-field calls can therefore multiplex well beyond the 33 samples used here, whereas four-field typing and full-gene reconstruction benefit from the depth that our current design provides, though future bioinformatic improvements could close this gap.

Benchmarking NGS-based HLA typing methodologies is inherently challenging because accuracy depends on the entire workflow, from raw sequencing read generation to the algorithms that produce HLA allele calls. The cause of incorrect HLA allele calls is not always immediately apparent, as errors may arise from data quality, the typing algorithm, or both. A meaningful comparison of HLA typing tools must therefore account for the sequencing platform, its error profile, and the library strategy, as WGS, whole-exome sequencing (WES), amplicon, and hybrid capture libraries may yield fundamentally different coverage profiles across HLA genes. HLA typing algorithms also vary in their robustness to coverage depth and in their accuracy across loci, leading to situations in which the highest-performing tool depends on both the gene and the depth of coverage^86^. This variability is expected given the substantial differences among HLA genes in size, polymorphism, and repeat content. Our own downsampling analysis showed different coverage requirements for the different HLA genes. For these reasons, the HLA allele concordance values presented here should be interpreted as specific to the high-coverage target-capture data generated with our protocol. In this context, HLA-Resolve outperformed all three tools we benchmarked on our PacBio sequencing reads.

The benchmarking datasets used to develop and validate open-source HLA typing tools have important limitations. The most commonly used benchmark, including for long-read typers SpecImmune^47^ and StarPhase^46^, is the 1KGP HLA allele labels^87^, which were generated from short-read whole-exome sequencing data using the PolyPheMe HLA typing tool (Xegen, France) at two-field resolution, limiting how finely any tool benchmarked against these labels can be evaluated. Furthermore, these labels have not been independently confirmed with newer sequencing and HLA typing technologies, and coverage depth varies considerably across the Phase 3 samples used to generate them^86^. Although ONT data are now freely available for many of the 1KGP samples, the 1KGP HLA allele labels have, to our knowledge, not been re-evaluated. More recently, Lai et al.^78^ generated allele labels for the HPRC v1 cohort by comparing the assembled sequences with alleles in the IPD-IMGT/HLA database. However, this approach assumes the assemblies are accurate across the HLA region. The generated allele labels do agree with the Immuannot annotation^88^ for the same assemblies at every allele where both are resolved, but both approaches evaluated the same underlying sequence. Any individual approach to establishing ground truth will have limitations, underscoring the need for consortium-oriented benchmarking datasets representing consensus across independent groups using different wet-lab and computational methods. The 19^th^ IHIWS reference cell line project is one such effort, to which we contributed data from our long-read hybrid capture method. Its consensus typings will allow an additional accuracy assessment of our method once they are released. Comprehensive long-read whole-genome characterization of the IHWG catalog would also provide a strong foundation for this effort, and is increasingly tractable as per-sample costs fall. Phased full-length allele sequences are also needed for HLA imputation reference panels, whose accuracy degrades substantially in populations underrepresented in existing panels^89^.

There is a rapidly growing interest in Oxford Nanopore Technologies (ONT)-based workflows because of the substantial advantages of long-read sequencing and the competitive pricing of the ONT MinION instruments^70,71^. These workflows represent a considerable improvement over older PCR-SSP and PCR-SSO methods, perform well at lower levels of resolution, and are beginning to enter clinical use. In our data, ONT small-variant genotype concordance in the HLA region was notably lower than PacBio, despite sequencing on the PromethION platform with R10 chemistry at higher average coverage depth. The effect on HLA typing was smaller and tool-dependent. Pooled across both cohorts, ONT concordance at three-and four-field resolutions was about 3 percentage points lower than PacBio for StarPhase, 2 percentage points lower for SpecImmune, and within 1 percentage point for FuFiHLA. Since ONT depth exceeded PacBio in 85% of sample-gene combinations, the gap may reflect the approximately 1.5x longer PacBio reads or ONT’s higher per-read error rate. We did not extend the same comparison to HLA-Resolve as it is currently PacBio-specific. Commercial typing software such as GenDx NGSengine or Omixon NanoTYPER may compensate further for ONT’s higher per-read error rate, but to our knowledge, no study has run these tools on matched PacBio and ONT data. Furthermore, even if commercial algorithms recover the correct allele label, it remains unclear how accurately they reconstruct full-length allele sequences, an important consideration for research in evolutionary genetics and immunogenetics.

In summary, we developed an open, end-to-end long-read HLA typing framework that combines a streamlined, automatable sequencing protocol with a fully integrated bioinformatics platform. Our results demonstrate that this approach delivers high-accuracy, high-resolution HLA allele calls across the classical HLA loci and reconstructs full-gene phased sequences. Our panel also extends comprehensive coverage to the MHC Class III region, which existing long-read HLA assays do not target. By making both the sequencing protocol and HLA-Resolve freely available, we provide a practical and cost-effective alternative to proprietary assays for targeted MHC sequencing and HLA typing.

## Methods

### Sample selection and cohort design

Thirty-three human cell line samples were selected for resequencing from the Coriell Institute/Human Pangenome Reference Consortium (HPRC) and International Histocompatibility Working Group (IHWG) catalogs (Supplementary Table 1) to enable three complementary benchmarking analyses: (i) genotype concordance with the Genome in a Bottle (GIAB) benchmark, (ii) HLA gene sequence reconstruction accuracy relative to the phased HPRC genome assemblies, and (iii) HLA typing concordance with the HPRC and IHWG reference typings. The HPRC cohort comprised 17 gDNA samples obtained from the NIGMS Human Genetic Cell Repository at the Coriell Institute for Medical Research, thirteen of which were included in the first HPRC pangenome reference release^43^. To support trio-based evaluation of genotype concordance and phasing accuracy, we additionally included the parents of HG002 (NA24149, NA24143; NIST IDs HG003, HG004) and HG005 (NA24694, NA24695; NIST IDs HG006, HG007), forming the two GIAB benchmark trios. The second cohort consisted of gDNA from 16 B-lymphoblastoid cell lines from the IHWG Cell and DNA Bank, each with previously curated HLA typings, enabling independent benchmarking of HLA allele typing performance. A third set of 96 samples, captured and sequenced after the main cohort for the 19th International HLA and Immunogenetics Workshop, was used to assess reproducibility of the capture assay (see “Hybrid capture reproducibility set”).

### Tagmentation and hybrid capture

Long-read target-capture libraries were generated using a custom barcoded tagmentation strategy, followed by hybrid capture enrichment of select MHC protein-coding genes. Custom barcoded transposase adapter oligonucleotides were designed for DNA tagmentation and adapter insertion. All oligonucleotides were ordered from Integrated DNA Technologies (IDT) and resuspended at 100 μM in IDTE (Supplementary Table 1). Barcoded oligonucleotides have a 5’ universal PCR adapter sequence (5’-cgaacatgtagctgactcaggtcac-3’) followed by a 16bp sample barcode and end with the transposase mosaic end binding sequence (5’-AGATGTGTATAAGAGACAG-3’). Partially double-stranded adapters were formed by annealing each barcoded oligonucleotide to a mosaic end reverse oligonucleotide ([5’-PHO]CTGTCTCTTATACACATCT) following the Diagenode Transposome Assembly protocol. Briefly, 9 μL each of 100 μM barcoded and mosaic end reverse oligonucleotide were mixed with 2 μL 0.5M NaCl in a 20 μL final volume. The reaction was incubated at 95°C for 5 minutes, cooled to 65°C at -0.1°C per second, incubated at 65°C for 5 minutes, and then cooled to 4°C at -0.1°C per second. Annealed adapters were stored at -20°C.

Barcoded transposomes were assembled by binding 2 μL unloaded tagmentase (C01070010-20, Diagenode) with 2 μL partially double-stranded adapter at 23°C for 30 minutes. A single barcoded adapter was used per tagmentation reaction, ensuring that both fragment ends carry the same barcode and enabling amplification with a universal PCR primer. Assembled transposomes were diluted in Tagmentase Dilution Buffer (Diagenode, C01070011) for storage and titrated to working concentrations to yield desired fragment sizes.

Genomic DNA was tagmented using the assembled barcoded transposomes. Tagmentation reactions containing 0.5 μg of genomic DNA, 17 μL of 2x tagmentation buffer (C01019043-1000, Diagenode), and 2 μL of diluted barcoded transposome, in a final volume of 34 μL, were incubated at 55°C for 15 minutes. 8.5 μL of 0.2% SDS was added to stop the reaction, and the reaction was incubated at 68°C for 5 minutes to inactivate the tagmentase. Low-molecular-weight DNA and unincorporated adapters were removed using 21 μL (0.5x) of AxyPrep MAG PCR cleanup beads (Axygen MAGPCRCL500). After elution from beads, barcoded samples were pooled by sample cohort (Coriell/HPRC, IHWG) for gap repair. 50 μL of pooled tagmented DNA was mixed with 50 μL of 2x Kapa HiFi polymerase and incubated at 72°C for 10 minutes, followed by another bead cleanup. Fragment size distributions were assessed using the Agilent Genomic ScreenTape and reagents (Agilent 5067-5365, 5067-5366).

Hybrid capture enrichment was performed for the HLA genes. A new probe set was designed in collaboration with Twist Bioscience, combining their existing probe panel targeting twelve HLA Class I (HLA-A, -B, -C) and II (HLA-DPA1, -DPB1, -DQA1, -DQA2, -DQB1, -DRB1, -DRB3, -DRB4, and -DRB5) genes (Twist_LRHLA_v3_T123plus; Twist Design ID: LRTE-97930942) with a novel probe set targeting the 69 Class III protein-coding genes (Twist Design ID: LRTE-96945019) annotated in the human reference genome. Hybrid capture was performed separately for the HPRC and IHWG pools. Tagmented, gap-repaired DNA was quantified with a Qubit DNA High Sensitivity kit (Thermo Fisher Q32851), and 3.5–4 μg of tagmented DNA was used directly as input for hybrid capture following Twist’s Long Read Library Preparation and Standard Hyb v2 Enrichment beginning at Step 4 (Preparing Libraries for Hybridization), except that a 100 μM Universal PCR Primer was substituted for Twist’s Universal Blockers. The Twist HLA Class I/II probe set and the new Class III probe set were combined at a 1:1 ratio for hybridization. Capture and wash steps were performed using the Twist Standard Hyb and Wash Kit v2 (Twist, 104446) with Dynabeads M-270 Streptavidin (Invitrogen, 65305), and post-capture amplification was carried out using KOD Xtreme Hot Start DNA polymerase (MilliporeSigma, 71975-3). We have authorized the vendor (Twist Bioscience) to make the full set of probes available for research use.

### Long-read sequencing, read post-processing, and coverage assessment

The PacBio library was prepared for sequencing following PacBio’s Preparing Target Enrichment Libraries for PacBio Sequencing using the SMRTbell Prep Kit 3.0 (PacBio, 102-141-700), with 150 ng of each post-capture amplification product as input. The library was sequenced on a Revio instrument at PacBio (Menlo Park, CA), with a movie length of 1,440 minutes (base caller v5.0). The ONT library was prepared using ONT’s Ligation Sequencing Kit V14 (SQK-LSK114) with 250–275 ng of each post-capture amplification product as input and sequenced on an ONT PromethION R10.4.1 flow cell at the UC Santa Cruz Sequencing Technology Center.

For the PacBio data, circular consensus sequences (HiFi reads) were generated using ccs v8.0.1, quality-trimmed with trim v1.1.0, and demultiplexed and adapter-trimmed with lima v2.10.0. PCR duplicates were removed before reference genome alignment using pbmarkdup v1.2.0 (https://github.com/PacificBiosciences/pbmarkdup). For ONT, base calling, demultiplexing, and adapter trimming were performed with Dorado v0.7.2+9ac85c6 (https://github.com/nanoporetech/dorado). For both platforms, the transposase mosaic end binding sequences from tagmentation were removed with cutadapt v5.1^90^. Reads were aligned to the GRCh38 reference genome (GCA_000001405.15; no-alt analysis set) using minimap2 v2.28-r1209^91^. ONT duplicates were marked after alignment using Picard MarkDuplicates v2.27.4^92^.

Sequencing coverage was assessed with mosdepth v0.3.10^93^, including only primary alignments with mapping quality of 20 or higher. For each sample, we computed mean coverage depth (Supplementary Tables 4–11) and the proportion of each feature covered by 20 or more reads (Supplementary Tables 8–11) for all genes, ncRNA genes, pseudogenes, and exons annotated in the HLA region of the GRCh38 GFF3 annotation file. Fold enrichment was calculated as the ratio of the mean on-target to the mean off-target depth. On-target depth was defined as the total number of target-aligned bases divided by the target region length (982,574 bp per the Twist Bioscience expected coverage BED file). Off-target depth was defined as the total number of non-target-aligned bases divided by the total off-target genome length, using a genome size of 3,099,922,541 bp (GRCh38 no-alt analysis set).

### Genotyping and phasing

Small variants were called with DeepVariant v1.6.1^94^ for PacBio and Clair3 v2.0.0 (model r1041_e82_400bps_sup_v500)^95^ for ONT. Both DeepVariant and Clair3 were evaluated on ONT reads, and Clair3 performed better. For the PacBio reads, earlier DeepVariant models (v1.6) performed better than newer versions (v1.8, v1.9) on our target capture data. Structural variants were called with pbsv (https://github.com/PacificBiosciences/pbsv) for PacBio and Sniffles2^96^ for ONT. Tandem repeats were genotyped from PacBio reads using TRGT v4.0.0^97^. Phasing was performed with HiPhase v1.5.0^44^ for PacBio and LongPhase v2.0.0^98^ for ONT. HiPhase outputs haplotype block coordinates directly, which were used to assess phasing performance in downstream analyses. For LongPhase, haplotype block coordinates were extracted from the phase set (PS) tags using WhatsHap stats v2.3^99^.

### Genotype concordance with Genome in a Bottle samples

Genotyping accuracy was assessed following the Global Alliance for Genomics and Health (GA4GH) best practices for benchmarking germline small-variant calls^80^. Genotype concordance was calculated against the GIAB high-confidence benchmark (v4.2.1) for samples HG002–HG007 across both sequencing platforms. The concordance analysis was restricted to genomic intervals with observed coverage to avoid misclassifying untargeted regions as false negatives. Specifically, we restricted the analysis to the 85 protein-coding HLA genes consistently captured by our probe set, using coordinates from the GRCh38 GFF3 annotation. Although targeted by our probe set, HLA-DRB3 and HLA-DRB4 were excluded as they are not part of the GRCh38 primary assembly. We adjusted the coordinates for HLA-B, HLA-F, and HLA-DQA1 to remove several small regions not targeted by the Twist probe panel, corresponding to exons beyond the canonical transcript boundaries (Supplementary Fig. 12a–c). We also excluded intron 9 of C4A and C4B, which contains a human endogenous retrovirus (HERV) insertion identical in sequence between the two paralogs that impedes alignment to the reference genome and results in coverage gaps (Supplementary Fig. 12d,e). In GRCh38, C4A and C4B differ at only 18 nucleotide sites, clustered in the region spanning exons 20–31. Reads originating from intron 9 rarely extended into unique flanking sequence and therefore mapped ambiguously to both paralogs, resulting in a mapping quality (mapQ) value of zero. These restrictions ensure that genotype concordance metrics reflect variant calling performance within confidently sequenced intervals. Genotype concordance was assessed with hap.py (https://github.com/Illumina/hap.py), using rtgtools vcfeval v3.12.1^100^ as the comparison engine.

We assessed the relationship between genotype concordance and coverage depth by iteratively downsampling PacBio reads from HG002. Due to differences in baseline coverage depth across the three HLA classes, each class was downsampled independently using Picard DownsampleSam v2.27.4^92^. For each downsample level, we computed the mean coverage depth of protein-coding genes, called variants with DeepVariant, and evaluated genotype concordance using hap.py. To account for stochastic variation introduced by random downsampling, we performed ten replicates per level and computed the mean and standard error for recall, precision, and F1 score.

### High-resolution HLA typing with HLA-Resolve

We developed HLA-Resolve, an HLA typing algorithm optimized for high-coverage PacBio reads. The pipeline takes raw PacBio reads in FASTQ or unmapped BAM format, reconstructs nucleotide sequences for each HLA gene through a conventional variant-discovery workflow, and queries the IPD-IMGT/HLA allele database to return best-guess HLA allele calls for eight classical HLA genes (HLA-A, -B, -C, -DPA1, -DPB1, -DQA1, -DQB1, -DRB1). The pipeline begins with optional adapter trimming with fastplong v0.2.2, the long-read implementation of fastp^101^, or cutadapt v5.1^90^, followed by alignment to a slightly modified version of the GRCh38 no-alt analysis set reference genome (GCA_000001405.15; see Supplementary Note 3 for details) with rammap v1.0.0^102^, a rust re-implementation of minimap2^91^. Reads placed in the HLA-DR region are then re-mapped competitively against a multi-allele HLA-DRB reference, and those whose primary alignment is not an HLA-DRB1 allele are removed (Supplementary Note 3). The alignment is then restricted to primary records in chr6:28,000,000-34,000,000. Genotyping is performed separately for each variant class: SNVs with bcftools v1.21^103^, indels with DeepVariant v1.6.1^94^, structural variants with pbsv v2.10.0 (https://github.com/PacificBiosciences/pbsv), and tandem repeats with TRGT v4.0.0^97^. Separate genotypers for SNVs and indels were chosen because DeepVariant commonly assigns RefCall (0/0 or ./.) genotypes when one haplotype is reference-like and the other is highly divergent, a pattern that mimics segmental duplication absent from the reference. In these cases, pileup-based genotypers proved sufficient given our high coverage depth. However, bcftools did not reliably genotype indels in our reads, so DeepVariant is used for indel calling, and HLA-Resolve implements a rescue function that promotes indel records from FILTER=RefCall to FILTER=PASS when genotypes show read depth (DP) >= 30 and alt allele depth (AD) >= 10. Rescued variants are classified as heterozygous if the variant allele frequency (VAF) falls between 0.3 and 0.7, or homozygous ALT if VAF exceeds 0.7. SNV genotypes from bcftools are filtered for DP >= 2, GQ >= 20, and QUAL >= 10. The SNV and indel calls are then concatenated into a single VCF, and SNV records that fall inside a homozygous deletion are removed.

The resulting genotypes are jointly phased with HiPhase v1.5.0^44^ and merged with bcftools concat. HLA-Resolve then filters redundant variant calls before sequence reconstruction. Because DeepVariant and pbsv may both genotype the same indels, and both may call variants at repeat loci better handled by TRGT, duplicate records must be removed before sequence reconstruction. HLA-Resolve’s filter first collects all pass-filter, phased structural variant genotypes from pbsv and records their genomic coordinates and affected haplotype. Each phased or homozygous indel that is not a pbsv structural variant (SV) is then checked for overlap with a collected SV on the same haplotype and suppressed if overlap is detected. An additional proximity check suppresses large indels (≥50 bp) within 10 bp of an SV start position, which likely represent the same calls due to left-alignment annotation differences. The SV overlap filter never suppresses SNVs, as they may represent true variants near SV breakpoints rather than redundant calls. A parallel filter is applied for TRGT, where non-TRGT variants falling entirely within annotated TRGT tandem repeat genotypes are suppressed, as the TRGT genotype already encodes the correct allele for the entire repeat span.

HLA-Resolve then inspects the haplotype blocks returned by HiPhase. Because haplotype block coordinates are defined by the first and last heterozygous genotype in a phase set, they exclude flanking homozygous bases. Following Holt et al.^44^, HLA-Resolve extends each block overlapping an HLA gene through adjacent homozygous regions up to, but not including, the next heterozygous genotype in either direction, then verifies that each gene is completely spanned by a single haplotype block before proceeding. Genes containing one or fewer heterozygous genotypes often do not fall within a defined phase set or haplotype block but are nonetheless effectively phased. When a single heterozygous genotype is present, allele assignment to each haplotype is arbitrary, and fully homozygous genes are identical on both haplotypes. Accordingly, genes with at most one heterozygous genotype are considered fully phased. Genes with internal phasing breaks follow a separate pipeline to extract the largest haplotype block possible for database querying (detailed below).

Before sequence reconstruction, coverage depth and breadth are evaluated for each target gene with mosdepth v0.3.10^93^, and genes that fail the minimum depth requirement are not reconstructed. For genes that meet coverage depth and phasing requirements, HLA-Resolve reconstructs full-length haploid nucleotide sequences by applying phased, pass-filter genotypes to the GRCh38 GFF3 gene models using a forked version of vcf2fasta^104^, maintained at https://github.com/matthewglasenapp/vcf2fasta and installed as a pip dependency pinned to release v1.0.0 (commit fa90bca). Full-gene intervals come from the Ensembl GRCh38.p14 gene records, and coding exons from the corresponding Matched Annotation from NCBI and EMBL-EBI (MANE) Select transcripts, with minor modifications for four genes. HLA-DQB1 is segregating for a splicing variant that skips exon 5, and the GRCh38 MANE Select transcript for HLA-DQB1 omits this exon, so we restored it, as HLA-Resolve performs germline HLA typing rather than transcript characterization. At three genes, the GRCh38 annotation carries substantially less untranslated sequence than the IPD-IMGT/HLA reference alleles do. We therefore padded the full-gene interval by 1kb at each end for HLA-C, and by 602 bp and 299 bp at the 3’ end for HLA-B and HLA-DPA1, respectively.

If a phasing break occurs within a gene, HLA-Resolve searches for the longest haplotype block that fully spans the antigen recognition site (ARS). When such a haplotype block exists, the region is rebuilt by rerunning vcf2fasta on a single-feature gene model truncated to the coordinates of the haplotype block. If no haplotype block fully spans the ARS, HLA-Resolve enters a rescue mode to extract as much information as possible (Supplementary Note 4). Even within defined haplotype blocks, a small fraction of genotypes may remain unphased, typically reflecting complex structural variants or low-confidence SNV genotypes. These unphased genotypes cannot be applied in a haplotype-aware manner and are therefore ignored, as are complex structural variant types such as breakend (BND), inversion (INV), and duplication (DUP), which are not currently compatible with tools like vcf2fasta and bcftools consensus. HLA-Resolve reports any such variants and any unphased genotypes that could not be applied.

Following allele sequence reconstruction, HLA-Resolve compares the reconstructed allele sequences to the alleles in the IPD-IMGT/HLA database using an iterative, hierarchical classification algorithm. Query and reference sequences are compared using edlib v1.3.9.post1^105^, which computes Levenshtein edit distance and returns the corresponding optimal alignment path as a CIGAR string. Edit distance is calculated in infix mode, in which leading and trailing gaps are not penalized, accommodating partial or incomplete reference sequences in the database as well as variable start and stop coordinates arising from different PCR primer designs. HLA-Resolve derives two additional classification metrics from the alignment path, named mismatch identity and match length. Mismatch identity is defined here as the proportion of bases that match at 1:1-aligned positions. We define match length as the total number of identical aligned nucleotide positions. In HLA-Resolve, database records are restricted to alleles whose feature annotation in the IPD-IMGT/HLA hla.xml file runs unbroken from the start of the gene, excluding alleles that were only partially sequenced. The program outputs a single best-match call for each haplotype, alongside a parallel file listing all equally scoring alleles where the call could not be resolved by the sequence.

HLA-Resolve first attempts to assign each reconstructed HLA allele sequence to a G group based on the antigen recognition site (ARS) sequence of the protein (exons 2 and 3 for HLA Class I; exon 2 for HLA Class II). G group reference sequences were derived from the allele exon annotations in the IPD-IMGT/HLA hla.xml database. Each G group ARS sequence is aligned against the query in infix mode. For Class I, the per-exon distances are summed. A G group is assigned only if each of its exons matches at zero edit distance. If a perfect match is identified, the corresponding G group is assigned, and a second search is performed using the full concatenated exon sequence, restricted to alleles within that G group. Candidate alleles are ranked by edit distance to provide the three-field allele assignment. Ties are broken by match length, then alphabetically. If no exact G group match is found, a full unrestricted search compares the concatenated exon query sequence against all alleles of the corresponding gene, using the same ranking and tiebreaking chain.

If the three-field allele assignment includes alleles with defined fourth-field values, a third search is performed using the full-gene reconstructed allele sequence (including introns and UTRs). Candidates are ranked by mismatch identity rather than by edit distance to avoid penalizing gaps due to unreliable intronic reconstruction. Ties are broken first by match length, favoring alleles with more aligned evidence, then by raw edit distance, and finally by lowest fourth-field value. Each tiebreaking tier is applied only to the candidates the previous tier could not separate. Using the lowest fourth-field value is effectively an allele-frequency prior, because fourth-field designations are assigned chronologically and earlier-annotated alleles tend to be the most common. We verified that HLA allele concordance was not inflated by the fourth-field tiebreaking chain (See Supplementary Note 5).

For the three genes HLA-DQA1, HLA-DQB1, and HLA-DRB1, HLA-Resolve implements a final re-consensus step. These genes often show large structural variants that can be difficult to genotype and phase with pbsv and HiPhase, often introducing phase breaks and requiring matching with a fragmented sequence. For a given gene, after the best-guess four-field allele calls are made, the tool extracts the mapped reads underlying those calls and assigns them to alleles. For genes with no pass-filter phased heterozygous genotypes, reads are pooled into a single consensus. For genes with phased heterozygous genotypes, reads are split by the HP tag assigned by HiPhase. To determine which HLA allele each HP tag corresponds to, reads carrying each tag are mapped against both best-guess HLA allele sequences, and each tag is assigned to the allele its reads match better. The two HP tag assignments are made jointly, so both tags cannot be assigned to the same allele. Reads whose primary alignment falls on the other HLA allele at a mapping quality (mapQ) of 30 or higher are discarded as mis-phased, and each remaining read set is re-aligned to its assigned allele with rammap to generate a consensus sequence using samtools consensus v1.21.0^106^ at default parameters. When both haplotypes share a three-field lineage, both consensus sequences are built against a single shared allele, so any difference between the two consensus sequences comes from the reads.

Each new consensus sequence is then re-matched to the database a final time, where the search is restricted to the four-field options within the same three-field family. Therefore, the re-consensus only refines the fourth field and never overturns calls at lower fields of resolution. The comparison uses only the exon and intron sequences, since database records vary widely in the amount of flanking untranslated sequence they include. When exon and intron sequences cannot separate two or more fourth-field candidates, the comparison is repeated over the full record with the untranslated regions trimmed to the span shared by all tied candidates, so that every candidate is scored over an identical interval. Re-consensus is rejected if (i) the database offers no fourth-field alternatives within the three-field group, or (ii) the re-consensus sequences are collectively a worse fit to their assigned alleles than the sequences they replaced. When both haplotypes share a three-field lineage and the reads are split by HP tag, the split is additionally compared against a single consensus built from all reads. The split consensus sequences are accepted only if their summed edit distance to their matched alleles is lower than twice the edit distance of the pooled consensus to its own best match. The pooled edit distance is doubled because a single pooled consensus represents both haplotypes. Positions where a consensus is ambiguous (i.e., “N”) count towards edit distance, because a pooled consensus is ambiguous at exactly the positions that distinguish two different alleles and would otherwise appear to match both candidates equally well. When a re-consensus is accepted, the re-consensus sequence replaces the existing sequence for that haplotype in the deposited gene and coding-sequence FASTA files, thereby recovering full-length, phased sequences for genes that phasing alone left fragmented.

### Validation of HLA-Resolve on hybrid capture libraries

HPRC and IHWG reference HLA allele typings were used to benchmark HLA-Resolve v0.9.9 (IPD-IMGT/HLA v3.64.0) against three long-read-friendly HLA typing algorithms: StarPhase v2.1.0 (IPD-IMGT/HLA v3.64.0)^46^, SpecImmune v0.0.2 (IPD-IMGT/HLA v3.64.0)^47^, and FuFiHLA v0.2.4 (IPD-IMGT/HLA v3.64.0)^48^. We also ran HLA*LA v1.0.4 (IPD- IMGT/HLA release not reported)^24^, which returns G group calls and was therefore not scored. StarPhase, HLA*LA, and SpecImmune require a mapped BAM as input. For StarPhase and HLA*LA, reads were mapped to the GRCh38 no-alt analysis set (GCA_000001405.15_GRCh38_no_alt_analysis_set) obtained from NCBI. For SpecImmune, reads were mapped to the GRCh38 full analysis set (GCA_000001405.15_GRCh38_full_plus_hs38d1_analysis_set) as recommended in its documentation. For all three, adapters were trimmed with cutadapt, reads were aligned with minimap2, and PCR duplicates, secondary and supplementary alignments were removed with Picard MarkDuplicates and samtools. FuFiHLA requires FASTQ input, so we provided FASTQ with adapters trimmed and duplicates removed. Reference HLA typings for the IHWG cohort were obtained from the Fred Hutchinson Cancer Center (https://www.fredhutch.org/en/research/institutes-networks-ircs/international-histocompatibility-working-group.html) and were, in most cases, identical to those available in the IPD-IMGT/HLA database. A small number of conflicts were manually resolved (Supplementary Note 6). These edits did not affect concordance between the tested HLA typing algorithms because they occurred in cases where all algorithms returned identical calls. Reference typings for the HPRC samples were taken from Lai et al.^78^ Supplementary File 6. The final truth HLA allele labels used in benchmarking are available in Supplementary Table 24.

To verify the accuracy of the reconstructed allele sequences, we calculated the edit distance between the HLA-Resolve output and the corresponding gene regions in the HPRC phased genome assemblies. We downloaded the HPRC release 2 assemblies and their Comparative Annotation Toolkit (CAT) v1.3 gene annotations from the HPRC (https://github.com/human-pangenomics/hprc_intermediate_assembly). If an assembly or annotation was missing for a sample in the release dataset, we fell back to the HPRC release 1 assemblies and their GFF3 gene annotations, which were downloaded from Ensembl (https://projects.ensembl.org/hprc/). This only applied to two samples, HG002 and HG03492. Full-gene (start codon through stop codon, including introns) nucleotide sequences for the classical HLA genes were extracted from each assembly using bedtools getfasta (v2.31.1)^107^ with start and stop coordinates from the corresponding CAT or GFF3 annotation. Because HLA-Resolve’s phased sequences are unlabeled with respect to parental origin, we tested both possible pairings against the HPRC maternal and paternal assemblies and selected the one with the lower total gap-compressed edit distance. Edit distance was calculated between the HLA allele sequences produced by HLA-Resolve and their corresponding sequences in the HPRC assembly using edlib^105^. For each edlib alignment, we also computed a gap-compressed edit distance by parsing the CIGAR string, treating each mismatch individually, and treating contiguous insertion or deletion characters as a single event. At HLA-DQA1, HLA-DQB1, and HLA-DRB1, this comparison evaluates the re-consensus sequence rather than the vcf2fasta reconstruction wherever a re-consensus was accepted, since the re-consensus replaces the deposited sequence.

### Validation of HLA-Resolve on whole-genome sequencing libraries

To assess HLA-Resolve on WGS input, we typed 146 public whole-genome PacBio libraries from 130 samples: 43 HPRC release 1 samples with reference typings from Lai et al.^78^, and 87 1000 Genomes Project samples sequenced by the HPRC in later years, for which two-field HLA-A, -B, -C, -DQB1, and -DRB1 typings are available from the 1000 Genomes Project HLA panel (https://ftp.1000genomes.ebi.ac.uk/vol1/ftp/data_collections/HLA_types/20181129_HLA_types_full_1000_Genomes_Project_panel.txt). We obtained reads from the Human Pangenome Reference Consortium data repository hosted on the AWS Open Data registry (s3://human-pangenomics), under the working/HPRC and working/HPRC_PLUS collections. We also obtained two additional HG002 libraries from the Genome in a Bottle Consortium. Each sequencing library was typed separately. Samples with two libraries were typed and evaluated independently. We concatenated SMRT cells from the same library with samtools cat into one unaligned BAM, adding SMRT cells until the estimated genome-wide coverage depth reached 30x and omitting any cell that would take it past 40x. The read files used for each library are listed in Supplementary Table 25. We ran HLA-Resolve v0.9.9 with --platform pacbio --scheme WGS, using IPD-IMGT/HLA release 3.64.0. Coverage was measured with mosdepth over the eight classical HLA genes and is reported as the mean of the per-gene mean depths. HLA-Resolve currently types a gene only when its antigen recognition site reaches ≥ 8× mean coverage. Genes falling below this threshold do not return allele calls and contribute to call rate rather than discordance. HLA allele calls were compared to the reference typings at every field of resolution the reference provides. An allele was evaluated at a given field resolution only if the reference specified it to that resolution. Any allele calls that resolved to fewer fields than the reference were scored as discordant, since failing to reach an available resolution is a typing failure.

### Coverage requirements for HLA typing with hybrid capture libraries

We measured the minimum coverage depth needed for accurate HLA typing by downsampling aligned reads from each sample to 19 target depths from 10-200x and retyping at each. Within each sample, we downsampled each gene independently to control for variation in coverage depth across genes. Here, depth refers to the length-weighted mean coverage across the coding sequence (CDS) exons of the gene model, measured with mosdepth v0.3.10. We first measured the baseline depth of each gene, then extracted reads for each gene with samtools v1.21.0, keeping only primary, non-duplicate alignments, in windows padded by 10 kb (5 kb for the HLA-DQ genes) so that each window fully covered the interval HLA-Resolve reconstructs. Because the HLA-DPA1 and HLA-DPB1 gene models overlap substantially, we extracted and downsampled them as a single region but report them separately. We downsampled each region with Picard DownsampleSam v2.27.4 at a proportion equal to the target depth divided by its baseline depth, then merged the seven independently downsampled regions into a single BAM for typing with HLA-Resolve. We passed downsampled BAMs to HLA-Resolve at the variant calling step, skipping adapter trimming, duplicate marking, alignment, and read filtering. We ran HLA-Resolve with its minimum-depth gates disabled. To control for variation from random subsampling, we performed ten replicates per target depth, each with a fixed random seed applied identically across genes, depths, and samples. We then calculated realized coverage depth for each gene, which is reported throughout in place of target depth. Because the HLA-DPA1 and HLA-DPB1 coding depths consistently differed, a shared downsampling level showed higher realized depth at HLA-DPA1 than at HLA-DPB1. For example, at the target depth of 200x, HLA-DPA1 showed 257x realized depth while HLA-DPB1 shows 142x. We plot realized depth in Supplementary Figs. 9-10, so HLA-DPB1 spans a narrower range and contributes two fewer levels.

We evaluated HLA allele concordance by gene and coverage depth. We restricted to 21 samples with sufficient depth to reach 200x across all seven downsampling regions, and with no evidence of contamination. We scored concordance per allele using the same rules as the full-depth benchmark, computed within each replicate by pooling the samples. We scored genes that could not be typed as discordant. We then averaged concordance across the ten replicates of each target depth and plotted it by realized depth, with error bars showing the standard deviation. We calculated the full-depth ceiling for each gene as HLA allele concordance recomputed on the same 21 samples with all reads.

We also evaluated gene sequence reconstruction accuracy as the edit distance between each downsampled reconstruction and the full-depth reconstruction of the same sample, computed with edlib v1.3.9 in infix (HW) mode. This comparison requires no reference allele typing and was evaluated on 25 samples with sufficient depth at every region and no evidence of contamination. We calculated gap-compressed edit distances, counting each indel run once. To determine the correct pairing between the downsampled and full-depth reconstructions, we tried both possible pairings and selected the one that minimized total edit distance. We compared four interval types: (i) the coding sequence, (ii) the span from the start codon to the stop codon with introns, (iii) the annotated transcript, and (iv) the full annotated Ensembl gene record. For each gene and depth, we report the proportion of reconstructions identical to the full-depth reconstruction and the median edit distance, pooled over samples and replicates. Because gene sequence reconstructions may become truncated at low depth due to phasing interruptions, we also measured sequence length and calculated the proportion of truncated reconstructions. Gene sequences with leading or trailing Ns were treated as truncated, with record length measured following the removal of flanking Ns, and the reconstruction was excluded from the edit-distance comparison. Ambiguous N bases inside the sequence were retained and scored as mismatches. We report truncation only for the coding sequence. We do not report truncation at the Ensembl gene record level because, for HLA-DQA1, -DQB1, and -DRB1, the length is set by the IPD allele used in the re-consensus step of HLA-Resolve, which may vary between runs, so no single truncation criterion applies across all eight genes. Furthermore, the transcript and start-stop intervals are derived from this gene interval, and here, truncation would only indicate whether the whole-gene reconstruction contained them.

### Hybrid capture reproducibility set

We prepared 96 additional samples for the 19^th^ International HLA and Immunogenetics Workshop (IHIWS) project “NGS of full-length HLA genes of reference cell lines.” This set consisted of 94 blinded gDNA samples from two quality-control and four unknown panels from the IHWG B-lymphoblastoid cell line collection (Fred Hutchinson Cancer Center), and two samples from the main cohort as inter-run replicates (HG002, IHW09224). Samples underwent tagmentation, gap repair, and hybrid capture as described above. For this set, samples were pooled by equal volume into eight pools of 12 samples. Hybrid capture was performed in three batches: pools 1–2, pools 3–4, and pools 5–8. Post-capture libraries were pooled into two sets of four pools (48 samples each) and sequenced on one PacBio Revio SMRT Cell per set at the UC Davis DNA Technologies Core. HiFi reads were demultiplexed with lima and processed as above. Pool-level quality was assessed per sample from unique mapped reads, target specificity, and fold enrichment (Supplementary Table 18). HLA typing was performed with HLA-Resolve v0.9.9 with IPD-IMGT/HLA 3.64.0 as detailed above. The sample codes and workshop consensus typings had not been released at the time of writing.

## Data Availability

Raw sequencing reads are available through the NCBI Sequence Read Archive (SRA) under BioProject PRJNA1417783. Coriell samples are deposited as runs SRR37075979 to SRR37076010, and IHWG samples as runs SRR40178127 to SRR40178158.

## Code Availability

HLA-Resolve is available at https://github.com/matthewglasenapp/hla_resolve. The code supporting the findings of this study is available on Dryad (DOI: 10.5061/dryad.sf7m0cgn0).

## Acknowledgments

We thank the following individuals for their contributions to this work. Arthur Chung, Casey Riegler, and Rebecca Barnard of Twist Bioscience developed the new Class III hybrid capture probe panel. PacBio sequencing was supported by Noelle Bittner, Ian McLaughlin, and Sarah Kingan, with additional bioinformatics support provided by Matthew Seetin and Xiao Chen. Brandy McNulty and Ivo Violich of the UC Santa Cruz Sequencing Technology Center assisted with nanopore sequencing and basecalling, and Jonathan Foos, Nicholas Scales, and Chris Still of Oxford Nanopore Technologies provided flow cells, sequencing kits, and technical support. Computational analyses were performed on the Hummingbird computational cluster at UC Santa Cruz, and we thank Rion Parsons and Jeffrey Weekley for system administration and technical support.

## Competing Interests

O.E.C., M.-C.Y., O.A.G., and M.R.G. are inventors on an international patent application filed by the Regents of the University of California through the UCSC Office of Technology Licensing. The application covers an assay for targeted long-read genomic sequencing of the human major histocompatibility complex.

Application number: PCT/US2026/044889

Filing date: September 9, 2026

Title: Methods of Genotyping a Genomic Region of a Subject

Inventors: Omar E. Cornejo, Muh-Ching Yee, Obed A. Garcia, Matthew R. Glasenapp

Priority: U.S. Provisional Application No. 63/879,493, filed September 10, 2025

## Declaration of generative AI and AI-assisted technologies in the manuscript preparation process

During the preparation of this work, the authors used Claude (Anthropic) to edit and refine manuscript text and to assist with writing and reviewing analysis code. The authors reviewed and edited the output as needed and take full responsibility for the content of the published article.

## Supplementary Information

### Supplementary Note 1: Variation in sequencing depth and duplication rates between hybrid capture cohorts

In the ONT PromethION sequencing run, the IHWG samples received approximately 5 times as many reads as the HPRC samples, likely due to unequal pooling of the two capture reactions before ONT library preparation (Supplementary Table 3). This imbalance was not present in the PacBio data, where the IHWG-to-HPRC read ratio was approximately 1:1 (Supplementary Table 2). PCR duplication rates were consistent within each cohort but differed substantially between cohorts due to overamplification of the IHWG pool during post-capture PCR. The average duplication rate for the HPRC samples was 19% for PacBio and 14% for ONT, compared to 61% (PacBio) and 80% (ONT) for the IHWG samples (Supplementary Tables 2 and 3). For PacBio, the higher duplication rate in the IHWG cohort led to lower on-target depth than in the HPRC cohort. This difference was not observed in the ONT data, where the disproportionate share of sequencing output allocated to the IHWG cohort offset its elevated duplication rate. Additionally, the equal mixture of the two Twist probe panels resulted in three-to four-fold greater coverage depth for the twelve targeted Class I and II genes than for the 69 Class III genes on both platforms.

### Supplementary Note 2: Contamination in sample IHW09117

One sample, IHW09117, showed signatures of contamination and was removed from HLA typing analysis. This sample showed an abundance of tri-allelic sites, a pattern absent from all other samples. Manual inspection of read alignments in Integrative Genomics Viewer (IGV) revealed >2 distinct haplotypes with strong support at multiple HLA genes. We were unable to determine whether the contamination occurred at the Fred Hutchinson Cancer Center or during our procedure.

### Supplementary Note 3: Reference genome modifications for HLA-Resolve

Sequencing reads from paralogs of the classical HLA loci can mis-map and introduce spurious genotype calls. For example, reads originating from the pseudogene HLA-Y often map to HLA-A^47^. HLA-Y is absent from the GRCh38 no alt analysis set reference genome, and was recently localized to a polymorphic 60 kb insertion that also includes the newly described HLA-OLI^108^. HLA-Y shows the highest sequence similarity to HLA-A of the HLA Class I genes and pseudogenes (see Fig. 3 from Alexandrov et al.^108^). We verified the identity of HLA-Y reads by comparing them to the HLA-Y alleles in the IPD-IMGT/HLA database, and prevented them from mapping to HLA-A by appending the 60 kb insertion sequence containing HLA-Y and HLA-OLI from GenBank sample HG00735 (GenBank accession JAHBCG010000038.1) to the GRCh38 reference genome as an additional scaffold.

The HLA-DRB paralogs also show high sequence similarity and are commonly misassigned during reference genome alignment. Reads from HLA-DRB1*07 and HLA-DRB1*09 alleles commonly mis-map to GRCh38 HLA-DRB5, and reads from an HLA-DRB1*04:12 allele preferentially map to HLA-DRB6. We therefore hard-masked HLA-DRB5 and HLA-DRB6 in the reference genome used by HLA-Resolve, forcing these reads to map to HLA-DRB1. All reads mapped to HLA-DRB1 are then filtered to remove those potentially originating from other DRB paralogs, including the HLA-DRB5 and HLA-DRB6 reads that hard-masking redirects there. To identify these reads, we constructed a multi-allele reference containing 33 sequence records. Seventeen records are full-length genomic HLA-DRB allele sequences from the IPD-IMGT/HLA database, comprising thirteen HLA-DRB1 alleles (one representative per allele group), three HLA-DRB3 alleles, and one HLA-DRB4 allele. Five records are paralog sequences extracted from genome assemblies rather than from IPD-IMGT/HLA. HLA-DRB6, -DRB7, - DRB8, and -DRB9 have no allele records in IPD-IMGT/HLA release 3.64.0, so HLA-DRB6 and HLA-DRB9 were taken from the GRCh38 primary assembly and HLA-DRB7 and HLA-DRB8 from the SSTO alternate haplotype. HLA-DRB5 was also taken from the GRCh38 primary assembly. The remaining eleven records are DRB-like intervals extracted from three HPRC assemblies (HG005, HG01928, and HG02630), covering additional HLA-DRB5 copies, RNU1-61P-adjacent segments, and intergenic regions carrying DRB-like fragments that may have arisen from partial gene duplication. An early step in the HLA-Resolve workflow re-maps reads that align to the HLA-DR region in GRCh38 against this reference to identify and remove those with primary alignments to anything other than HLA-DRB1 alleles.

### Supplementary Note 4: HLA-Resolve haplotype block recovery logic

Low variant density can interfere with read-based phasing and haplotype block assignment. When no single haplotype block fully spans the antigen recognition site (ARS) sequence of an HLA gene, HLA-Resolve attempts to recover as much information as possible by evaluating the distribution of heterozygous genotypes along the gene model. These edge cases are hypothetical and did not occur in our data, but could arise when one or more haplotype blocks overlap a gene and none completely spans the ARS, or when no haplotype block overlaps the gene and it carries more than one unphased heterozygous genotype. For example, if no haplotype block spanned the ARS of one of a sample’s HLA genes, but its exons were fully homozygous, and the unphased heterozygous genotypes fell only in introns/UTRs, accurate coding sequences could still be reconstructed. Furthermore, if unphased heterozygous sites occurred only in noncoding or non-ARS exons, an accurate ARS could still be reconstructed and used for HLA typing at G group resolution. To implement this rescue, heterozygous genotypes are counted separately across all CDS exons and across the subset that falls within the ARS. Depending on the distribution of heterozygous genotypes, HLA-Resolve does one of the following (documented in the HLA-Resolve GitHub under https://github.com/matthewglasenapp/hla_resolve/blob/main/docs/technical_reference.md):

a. If there are zero CDS heterozygous genotypes, the largest ARS-overlapping interval containing <= 1 heterozygous genotype is reconstructed by re-running vcf2fasta on a gene model truncated to that interval.
b. If exactly one heterozygous genotype is present across the entire CDS, a new interval is defined relative to the location of the heterozygous CDS genotype as (prev_het + 1) to (next_het -1). If this interval includes the full ARS, the gene record is reconstructed by rerunning vcf2fasta on a gene model truncated to that interval, which is then used for matching. If the interval does not fully span the full ARS, no gene record is produced, and allele matching stops after coding sequence matching (three-field resolution), and the allele does not receive a fourth-field assignment.
c. If more than one CDS heterozygous genotype exists, but at most one falls within the ARS CDS exons, only the ARS CDS subsequence is extracted, and the genomic sequence is extended outward from the ARS boundaries to the nearest flanking heterozygous genotypes. For HLA Class I genes under this condition, an additional check is performed for heterozygous genotypes in intron 2 (the intron separating the two ARS exons), and the genomic sequence is not reported if any are found. If more than one heterozygous genotype falls within the ARS CDS exons, the gene is excluded from typing.

### Supplementary Note 5: Performance of HLA-Resolve’s tiebreaking chain

After a three-field assignment is made, if the full-length reconstructed sequence cannot distinguish the remaining fourth-field candidates, HLA-Resolve assigns the fourth-field allele with the lowest value. That default is effectively an allele-frequency prior, because fourth-field designations are assigned chronologically and earlier-named alleles tend to be more common. To confirm that four-field concordance was not inflated by this rule, we examined how allele assignments were distributed across the tiebreaking chain and stratified concordance by fourth-field value. Across the 31 PacBio samples, 482 haplotypes received a fourth-field assignment. Sequence separated the candidates in 435 of them. The remaining 47 (9.8%) were assigned the lowest fourth field. Four-field concordance remained high (89%) when the truth allele’s fourth field was not :01 (140/157), compared to 92% when it was (175/191). Among the reference typings, :01 was the fourth field for 55% of truth alleles (191/348), which is expected given that the most common alleles tend to be discovered and named first.

### Supplementary Note 6: Resolution of Fred Hutchinson Cancer Center - IPD-IMGT/HLA conflicts

- IHW09122 HLA-C: IPD-IMGT/HLA shows C*07:02, C*02:02:02, while Fred Hutch shows C*07:02:01:03, C*15:02:01:01. We chose the Hutch typings because they are at higher resolution and supported by all the typing tools used in the benchmarking analysis.
- IHW09125 HLA-C: Both the IPD-IMGT/HLA and Fred Hutch show C*01. However, all typing algorithms tested, including our own, showed a heterozygous HLA type C*01/C*08 in both the PacBio and ONT data. Therefore, we edited the truth labels to C*01/C*08, but left the call at one-field resolution.
- IHW09359 HLA-DRB1: Fred Hutch has missing DRB1 HLA types, while IPD-IMGT/HLA shows DRB1*04 (homozygous). All HLA typing algorithms tested supported this call on both sequencing platforms, so we updated our truth set to include DRB1*04, DRB1*04.

For the remaining IPD-Hutch conflicts, we chose the HLA allele call made more recently or at higher resolution, which was likely to be typed more recently.

- IHW09198 HLA-DQB1: IPD shows DQB1*03:01:01, DQB1*05:02:01 while the Hutch shows DQB1*03:01:01:01/DQB1*03:01:01:03_NewIntronicVariant_hybridx1, DQB1*05:02:01
- IHW09224 HLA-DQA1: IPD shows DQA1*01:01:01:07, DQA1*01:02:01:05, while the Hutch shows DQA1*01:01:01:01/DQA1*01:01:01:02/DQA1*01:01:01:03, DQA1*01:02:01:01/DQA1*01:02:01:03/DQA1*01:02:01:05. Here, we chose the IPD typings, as they appear more recently updated.
- IHW09224 HLA-DQB1: IPD shows DQB1*05:01:24:01, DQB1*05:02:01:01, while the Hutch shows DQB1*05:01:01:03, DQB1*05:02:01. Here, we chose the IPD typings, as they appear more recently updated.
- IHW09251 HLA-DPB1: IPD shows DPB1*02:01:02, DPB1*04:02 while the Hutch shows DPB1*02:01:02|DPB1*416:01, DPB1*04:02:01:02|DPB1*105:01.

## Supplementary Figures

**Supplementary Figure 1:**
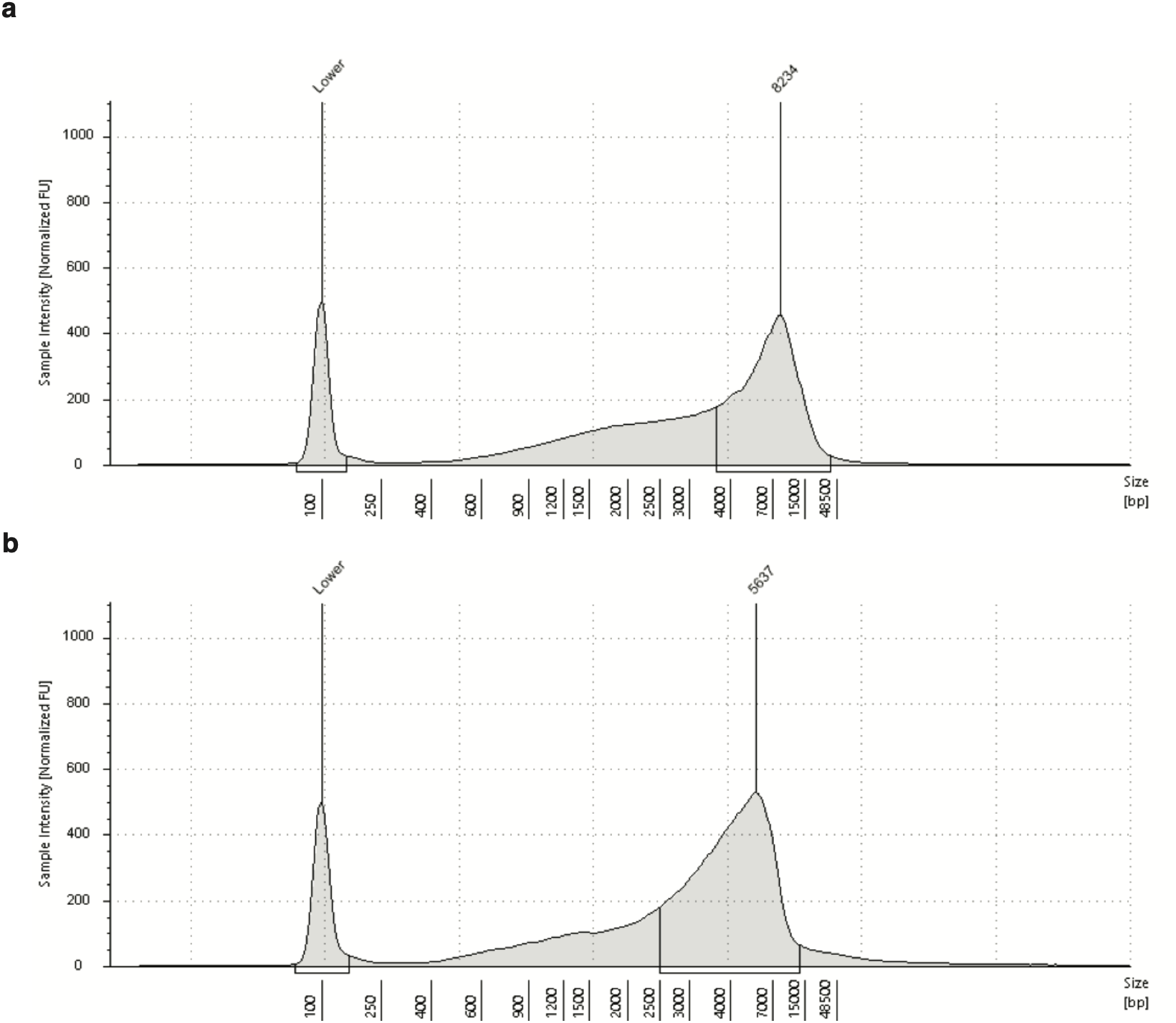
Distribution of DNA fragment lengths. Genomic DNA ScreenTape after tagmentation, before hybrid capture (**a**) and after hybrid capture and PCR amplification (**b**).

**Supplementary Figure 2:**
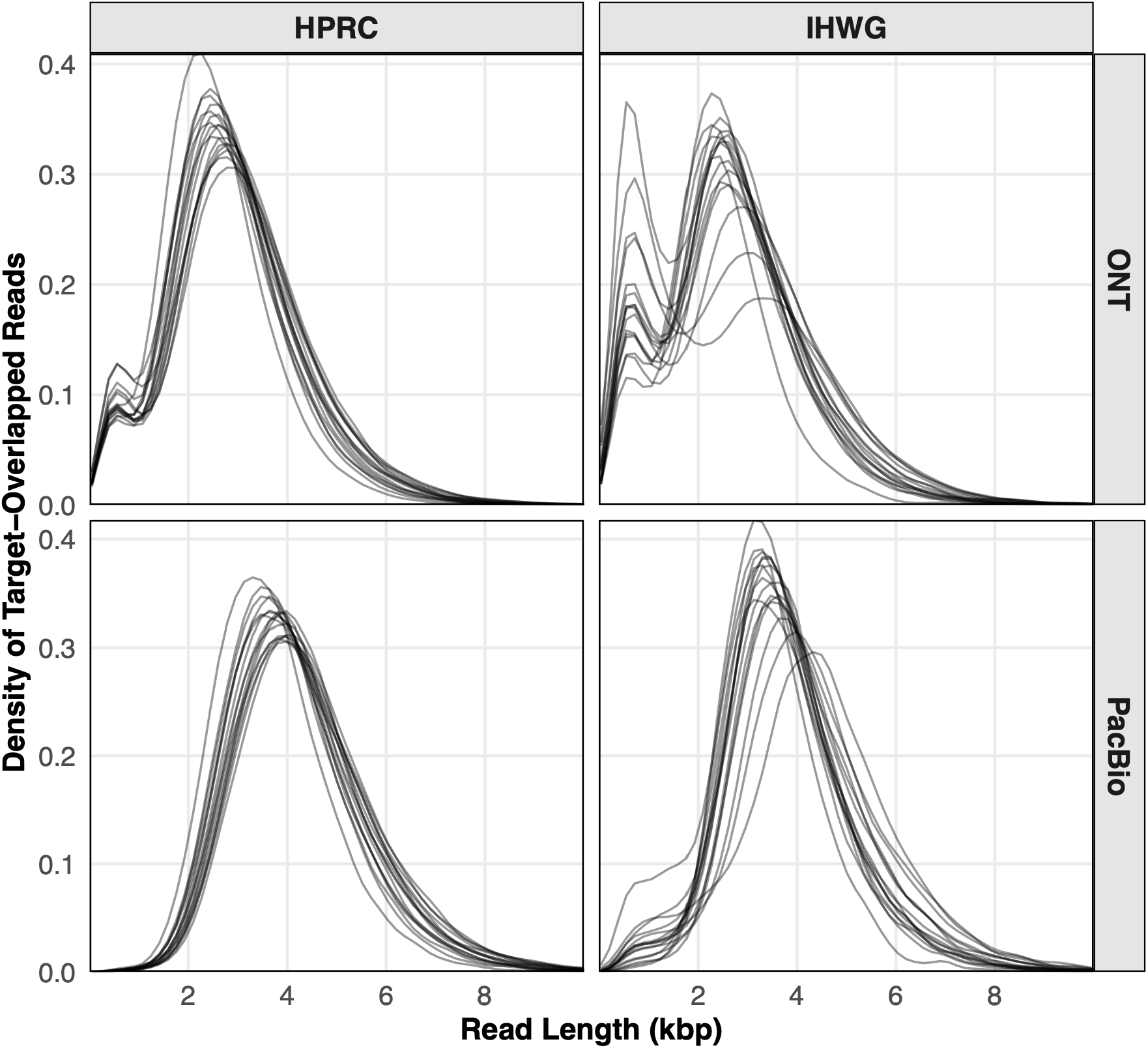
Distribution of sequencing read lengths for target-overlapped reads. Read-length distributions (in kilobases) for target-overlapped reads, stratified by sample cohort (HPRC, IHWG) and sequencing platform (ONT, PacBio). Each curve represents an individual sample.

**Supplementary Figure 3:**
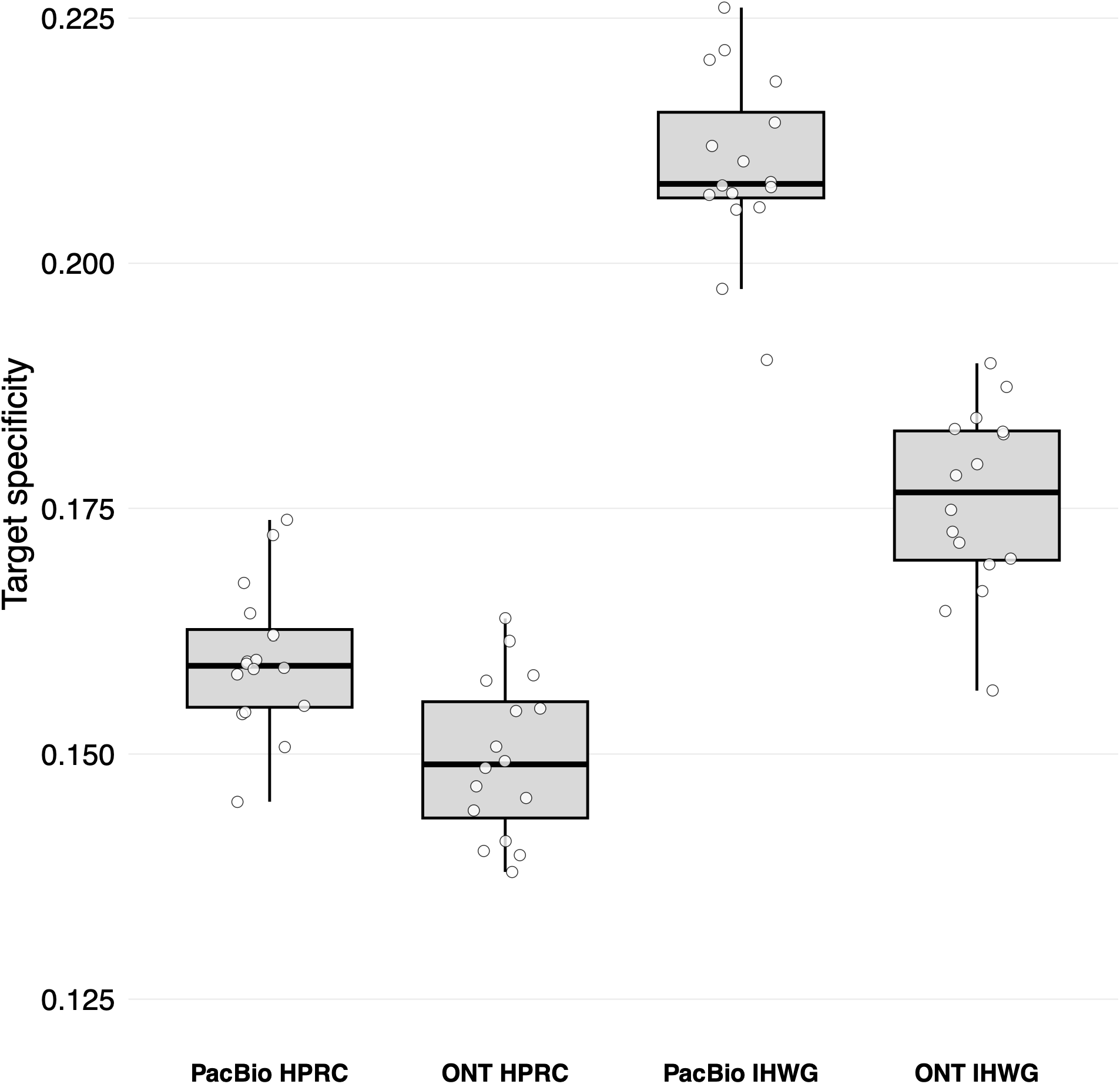
Target specificity by hybrid capture experiment and sequencing platform. Target specificity was calculated as the proportion of on-target aligned bases. Points represent individual samples.

**Supplementary Figure 4:**
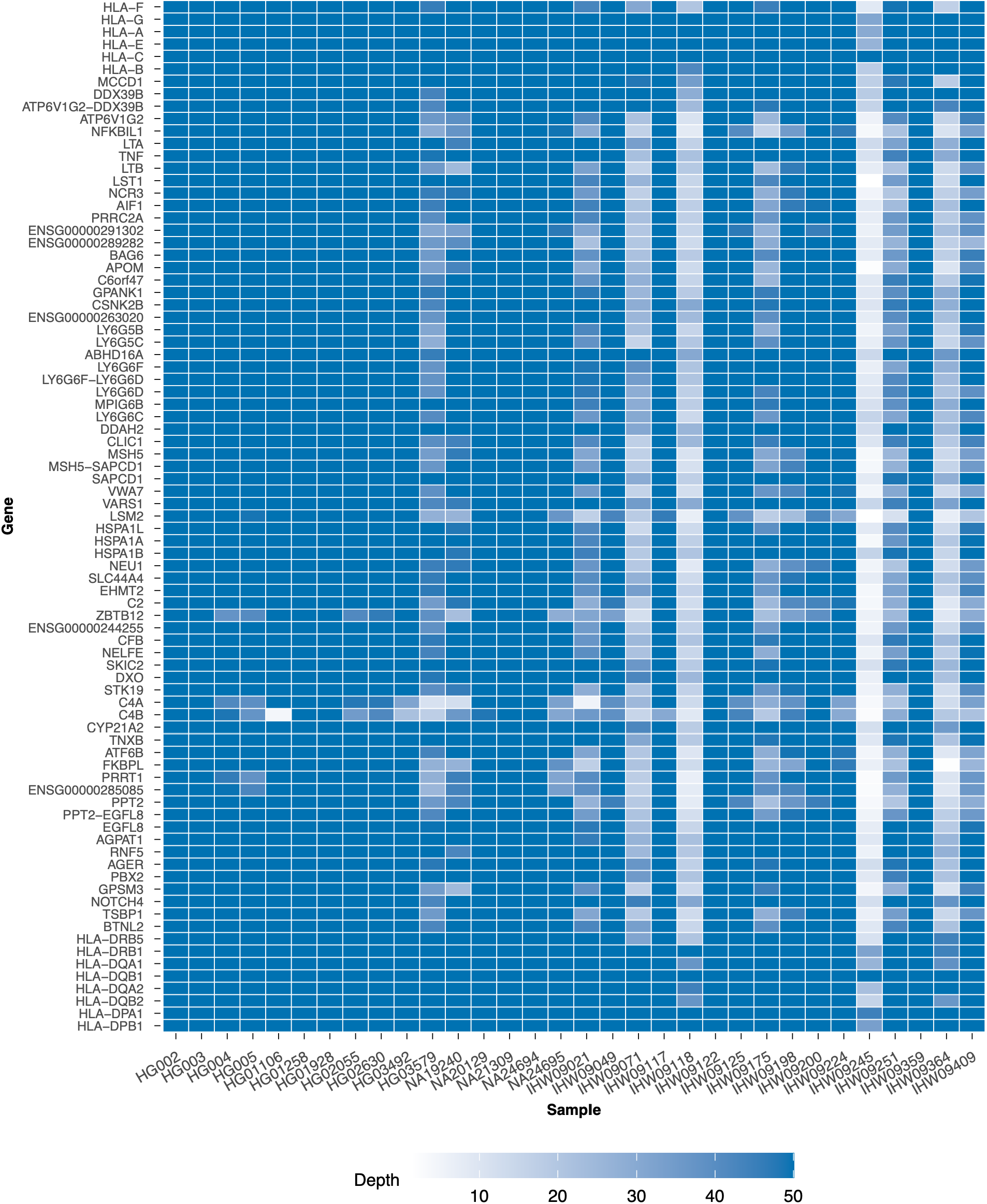
Gene-level PacBio Revio coverage across the captured HLA region. Heatmap showing mean sequencing depth for each captured protein-coding gene in the HLA region and each sample. Genes are ordered by genomic position along chromosome 6. Coverage depth is capped at 50x for display. All values above 50x are shown as 50x.

**Supplementary Figure 5:**
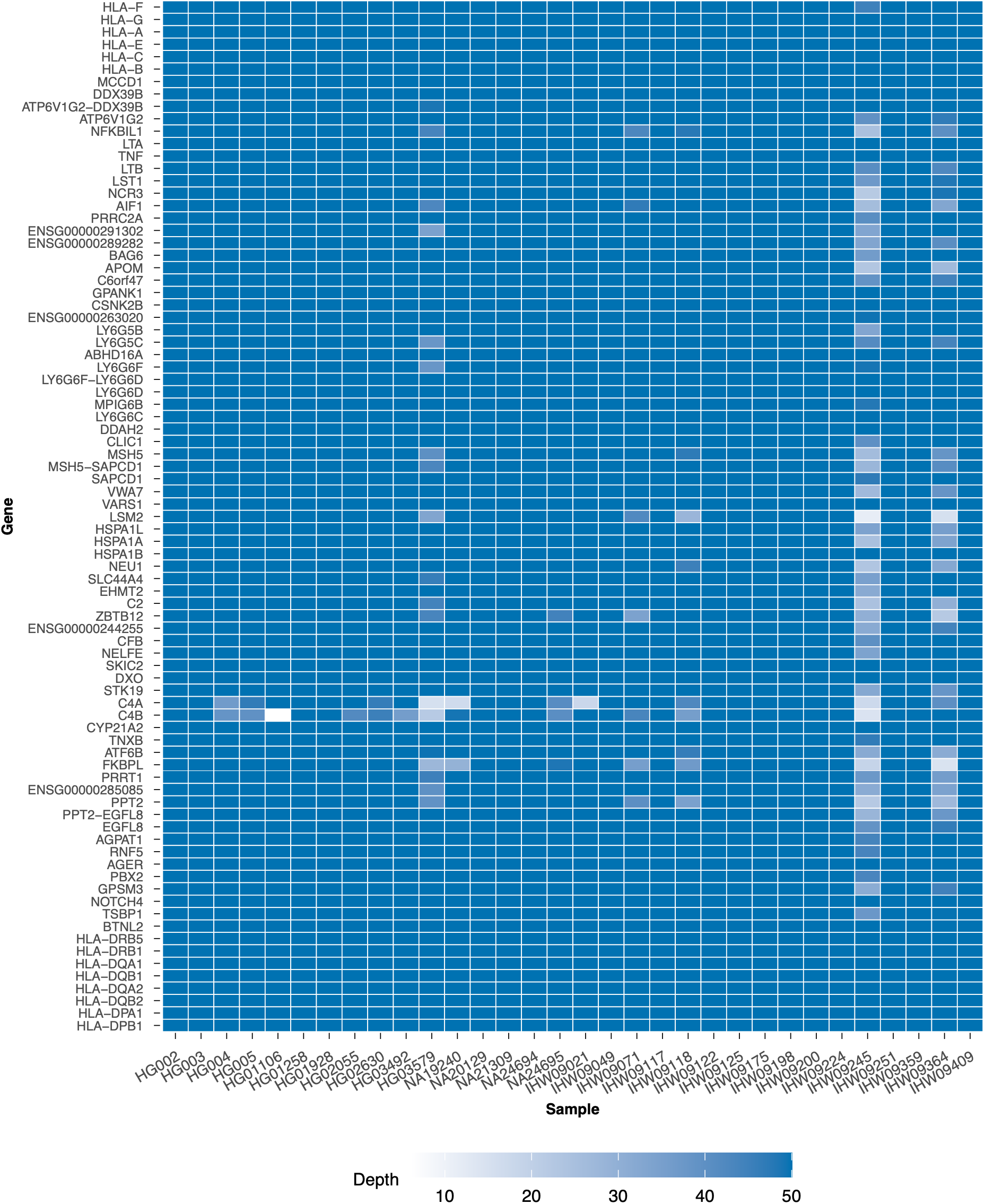
Gene-level ONT PromethION coverage across the captured HLA region. Heatmap showing mean sequencing depth for each captured protein-coding gene in the HLA region and each sample. Genes are ordered by genomic position along chromosome 6. Coverage depth is capped at 50x for display. All values above 50x are shown as 50x.

**Supplementary Figure 6:**
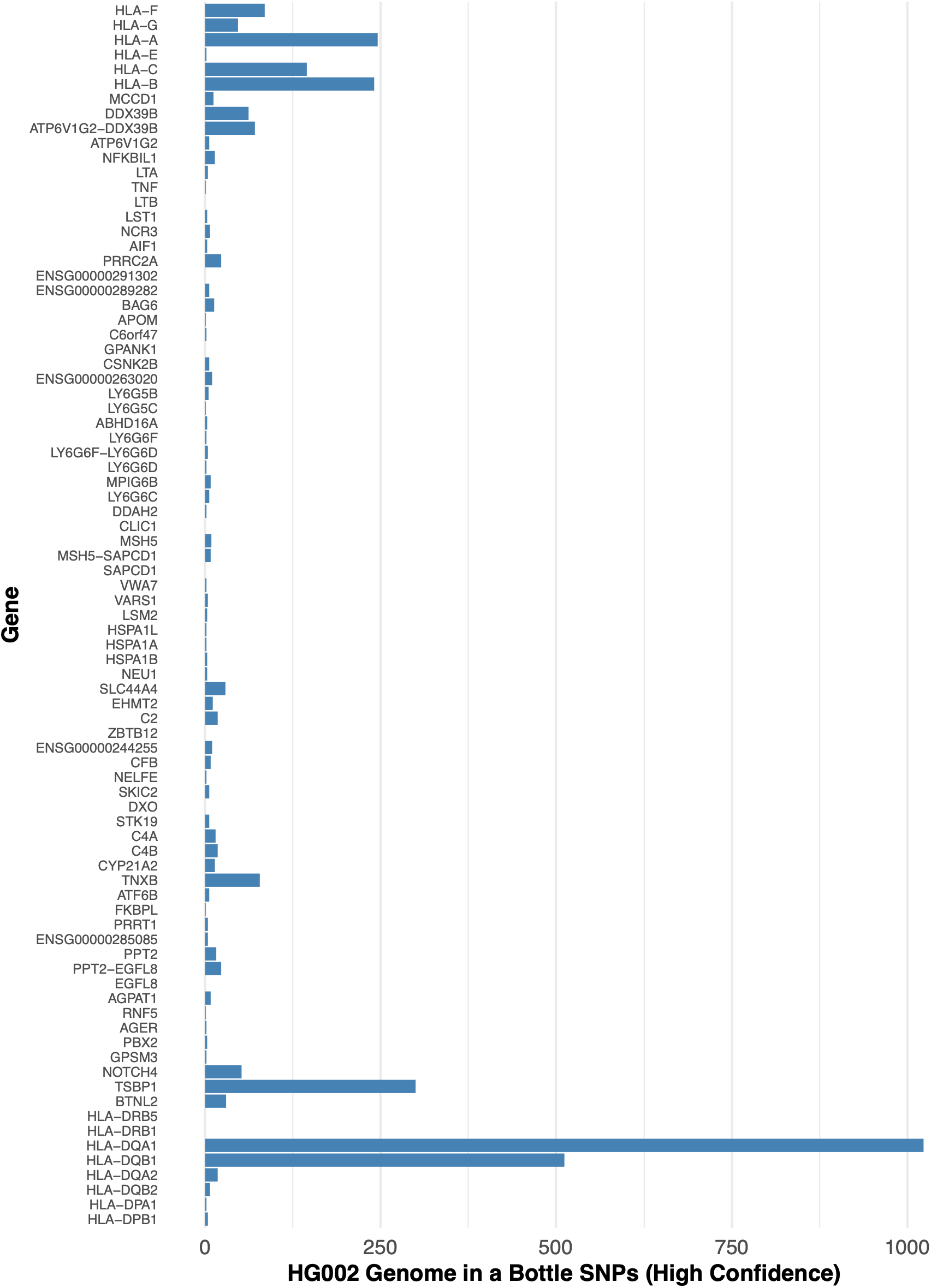
High-confidence Genome in a Bottle SNVs per captured gene in HG002. Number of high-confidence GIAB SNVs falling within each captured gene in the MHC region, ordered by genomic position. Counts total 3,312 across the 83 genes shown.

**Supplementary Figure 7:**
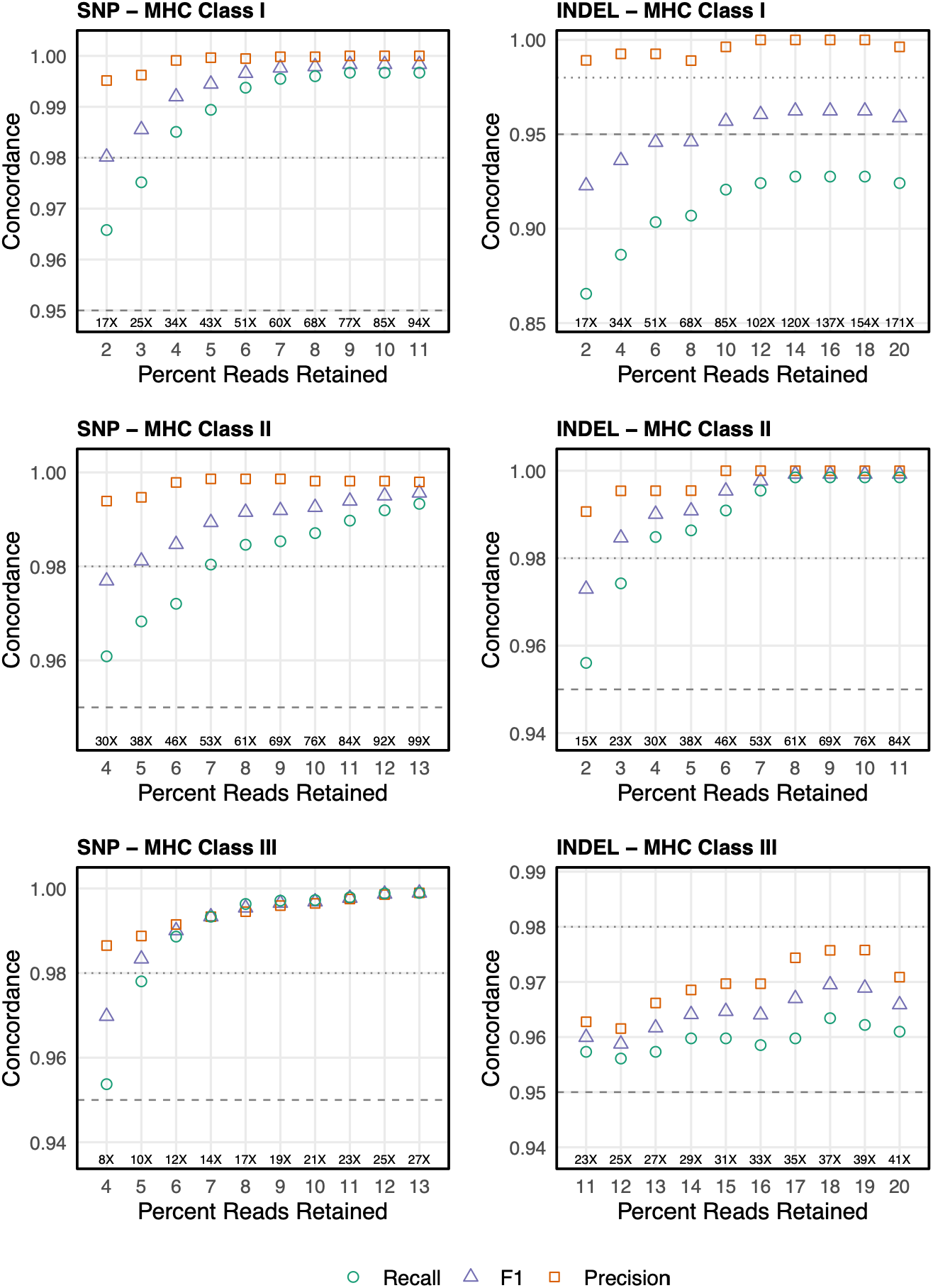
Effect of downsampling on HG002 genotype concordance. Recall, precision, and F1 for single-nucleotide variants (left) and indels (right) are shown for the Class I (top), Class II (middle), and Class III (bottom) regions across progressive levels of read downsampling. Metrics were computed from PacBio data after random read subsampling and genotyping with DeepVariant v1.6.1. The x-axis shows the percent of reads retained, with the resulting mean on-target coverage depth annotated. Genotype concordance with the Genome in a Bottle benchmark was assessed for variants within captured protein-coding genes using ten independent downsampling replicates. Points indicate the mean metric value across replicates.

**Supplementary Figure 8:**
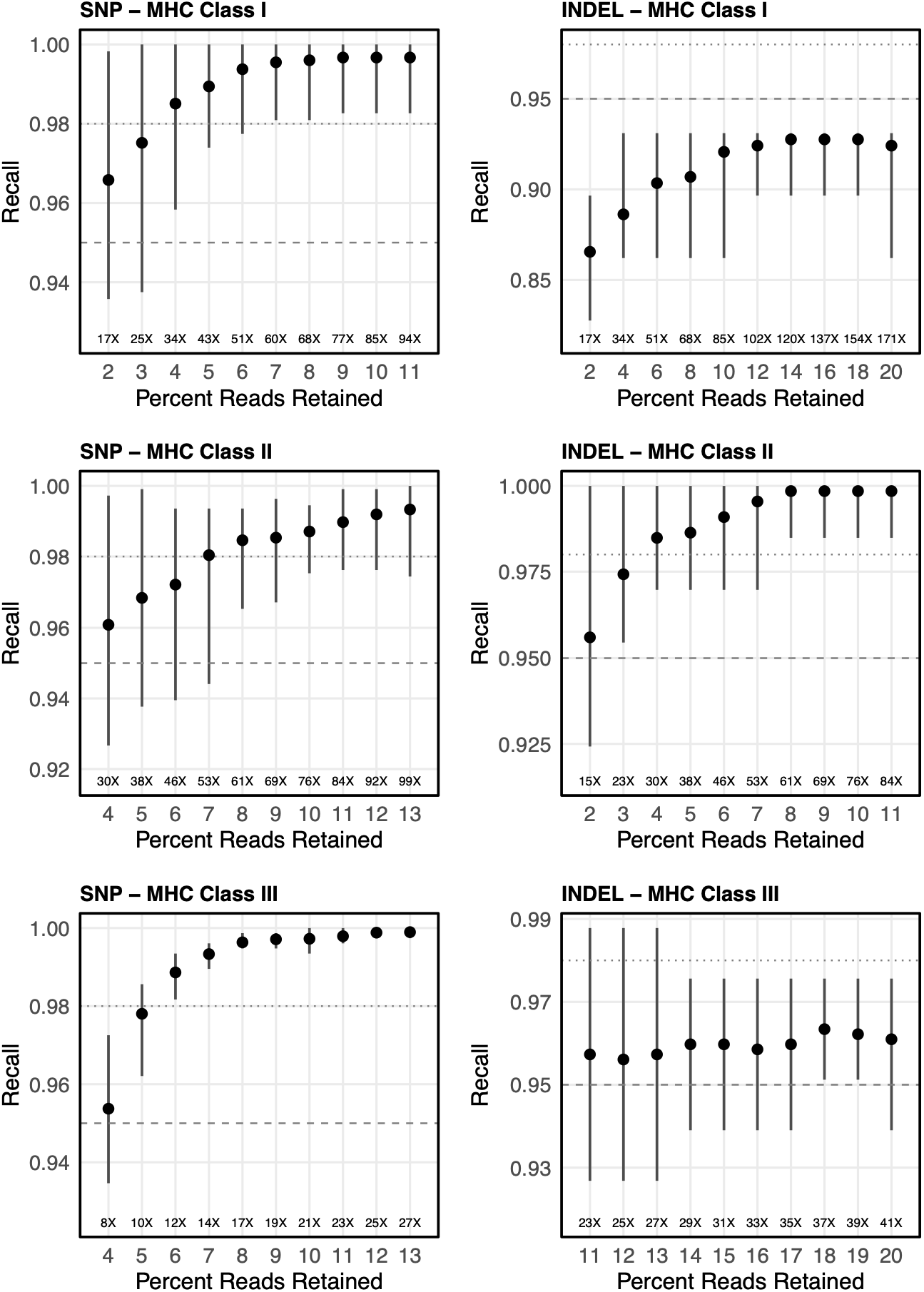
Effect of downsampling on HG002 genotype recall. Recall for single-nucleotide variants (left) and indels (right) is shown for the Class I (top), Class II (middle), and Class III (bottom) regions across increasing levels of read downsampling. Metrics were computed from PacBio data following random read subsampling and genotyping with DeepVariant v1.6.1. The x-axis shows the percent of reads retained, with the resulting mean on-target coverage depth annotated. Genotype concordance with the Genome in a Bottle benchmark was evaluated for genotypes within captured protein-coding genes across ten independent downsampling replicates. Points denote the mean recall across replicates, and vertical bars indicate the observed minimum-maximum range.

**Supplementary Figure 9:**
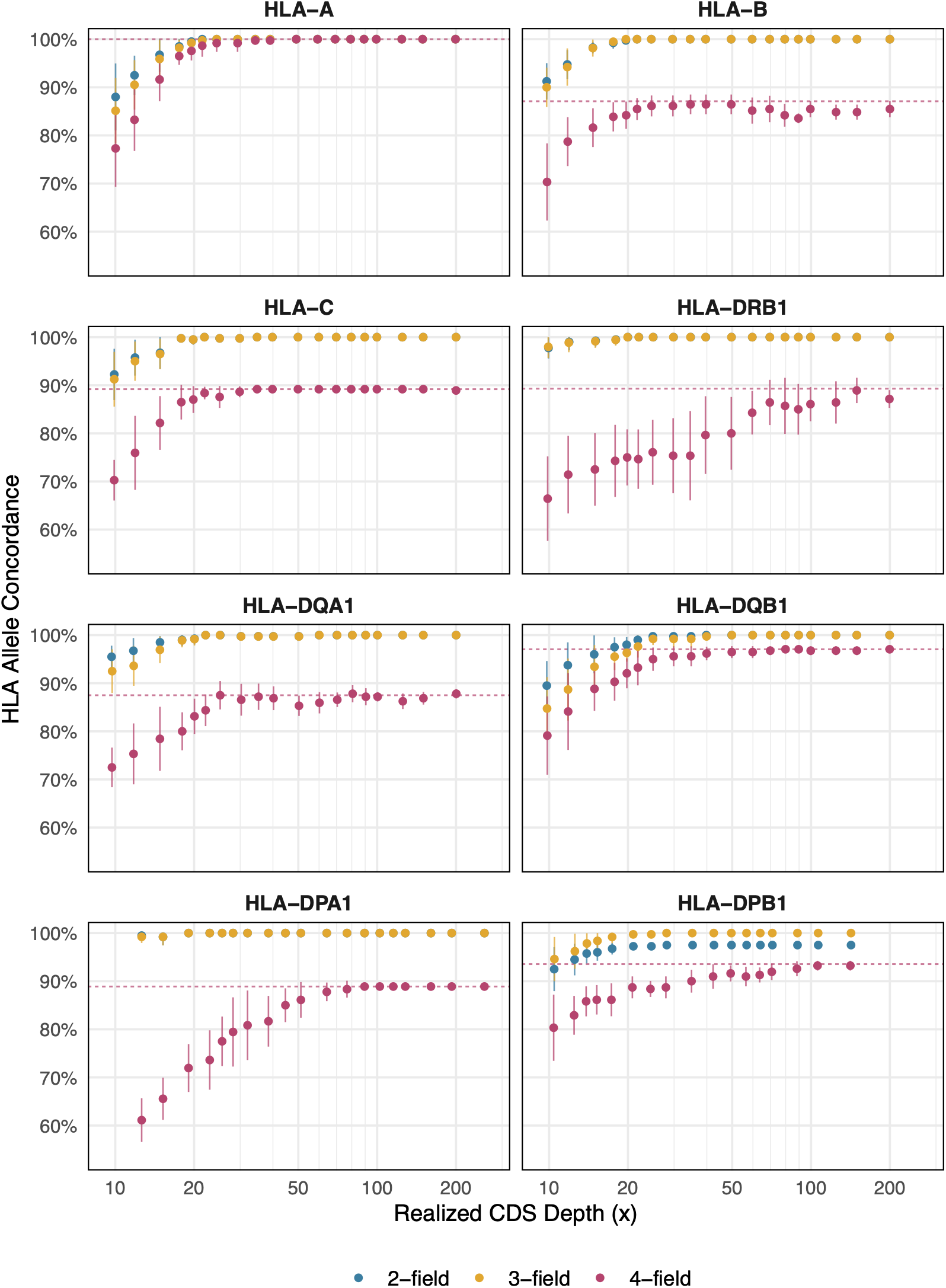
HLA typing concordance by coverage depth. HLA allele concordance at two-field (blue), three-field (gold), and four-field (magenta) resolution for each of the eight classical HLA genes, plotted against realized coding-sequence depth (log scale). Each gene was downsampled independently to 19 target depths, with ten replicates per depth. Points represent the mean concordance across replicates, with vertical bars showing the standard deviation. The red horizontal dashed line marks the four-field concordance value at full depth for the same samples, representing the typing ceiling.

**Supplementary Figure 10:**
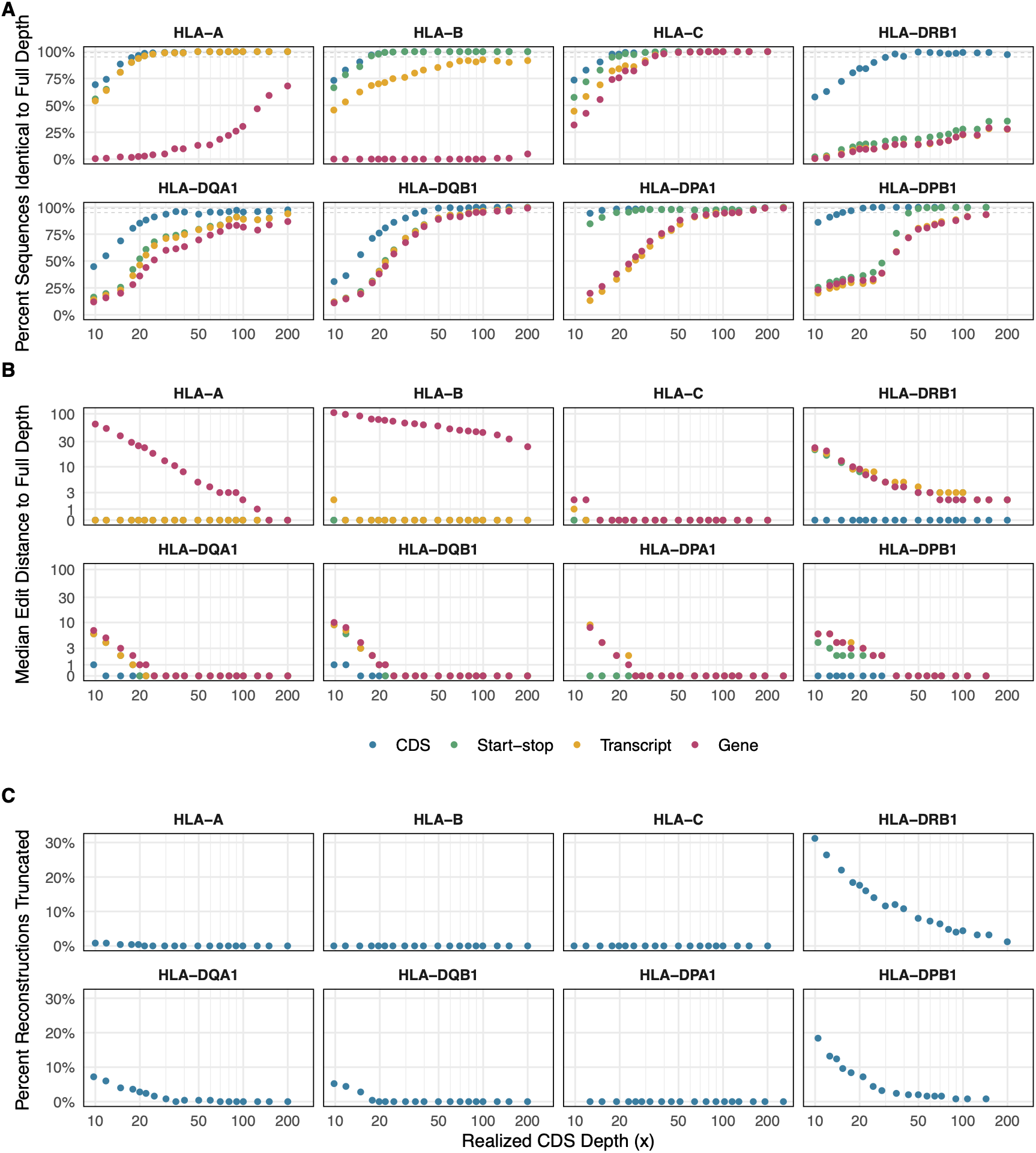
Gene sequence reconstruction accuracy by coverage depth. Each gene was downsampled independently to 19 target depths with ten replicates per depth, and each downsampled sequence reconstruction was compared to the full-depth reconstruction of the same sample using gap-compressed edit distance. Panels (a) and (b) show four intervals: the coding sequence (blue), the span from the start codon through the stop codon including introns (green), the annotated transcript (gold), and the full Ensembl gene annotation (magenta). (a) The percentage of sequence reconstructions identical to the full-depth reconstruction. (b) Median edit distance to the full-depth reconstruction. The y-axis is linear between 0 and 1 and logarithmic above, so reconstructions identical to full depth are shown at zero. (c) Percentage of coding sequence reconstructions shorter than the full-depth reconstruction. All x-axes show realized coding-sequence depth on a log scale.

**Supplementary Figure 11:**
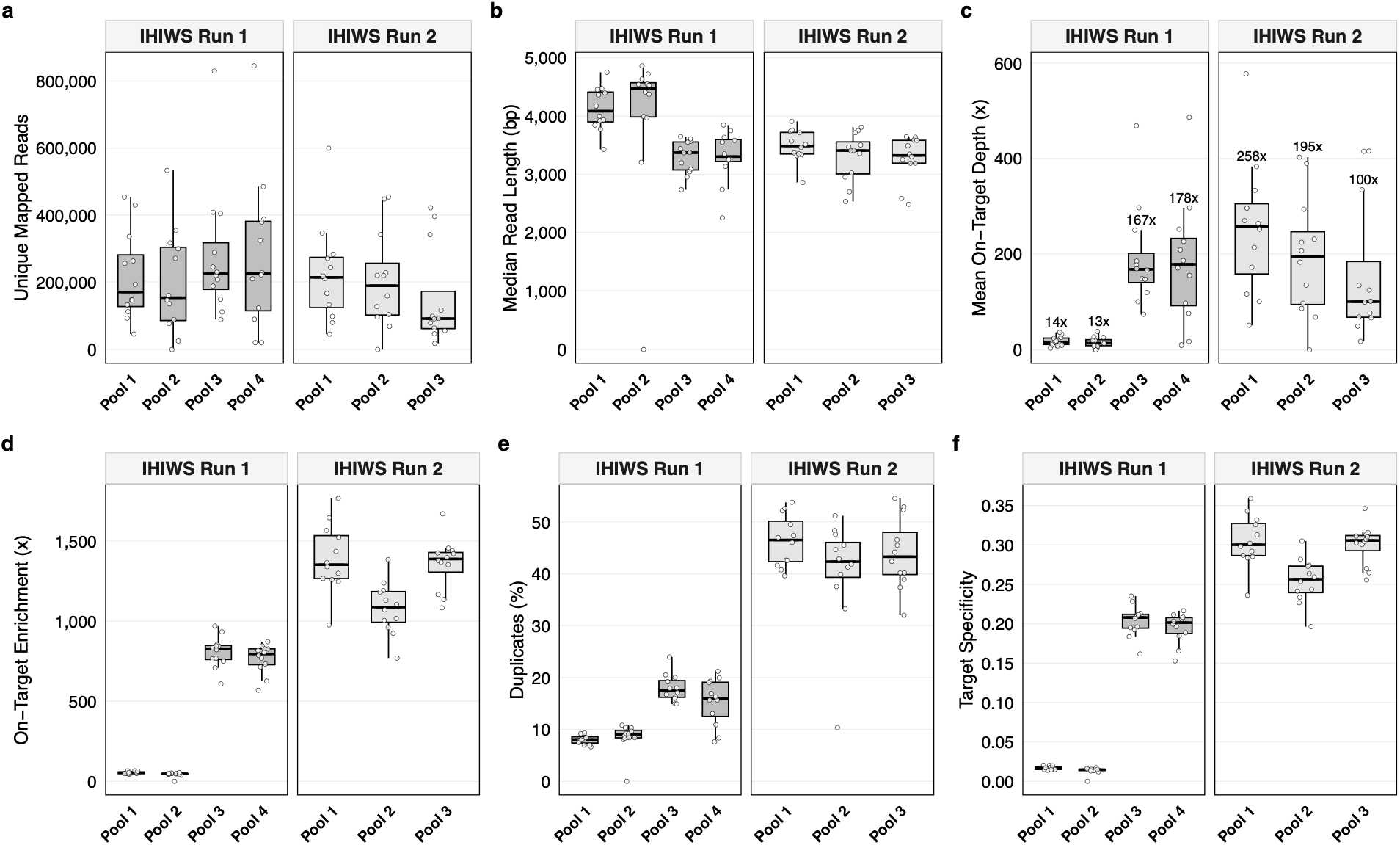
Summary of IHIWS Targeted Sequencing. **a**, Number of sequencing reads per sample, with PCR duplicates removed. **b**, Median length of target-overlapped reads. **c**, Mean on-target coverage depth for each sample, with the median for each pool labeled above its box. **d**, On-target enrichment, defined as the ratio of mean on-target to off-target coverage depth. **e,** PCR duplicate rate. **f**, Target specificity, defined as the proportion of aligned bases on target. In a-f, each point represents one sample. Boxes show the median and interquartile range, stratified by sequencing run and hybrid capture pool. Hybrid capture pool 4 of sequencing run 2 is omitted due to low library yield and insufficient reads per sample. The post-capture yield of hybrid capture pools 1 and 2 of sequencing run 1 was twice that of the other pools, consistent with off-target carryover during hybridization or washing. Although mean on-target depth in these pools was only 13-14x across the full capture target, median ARS depth at the eight classical HLA genes was 43-232x, sufficient for full-resolution HLA typing in 22 of the 24 samples.

**Supplementary Figure 12:**
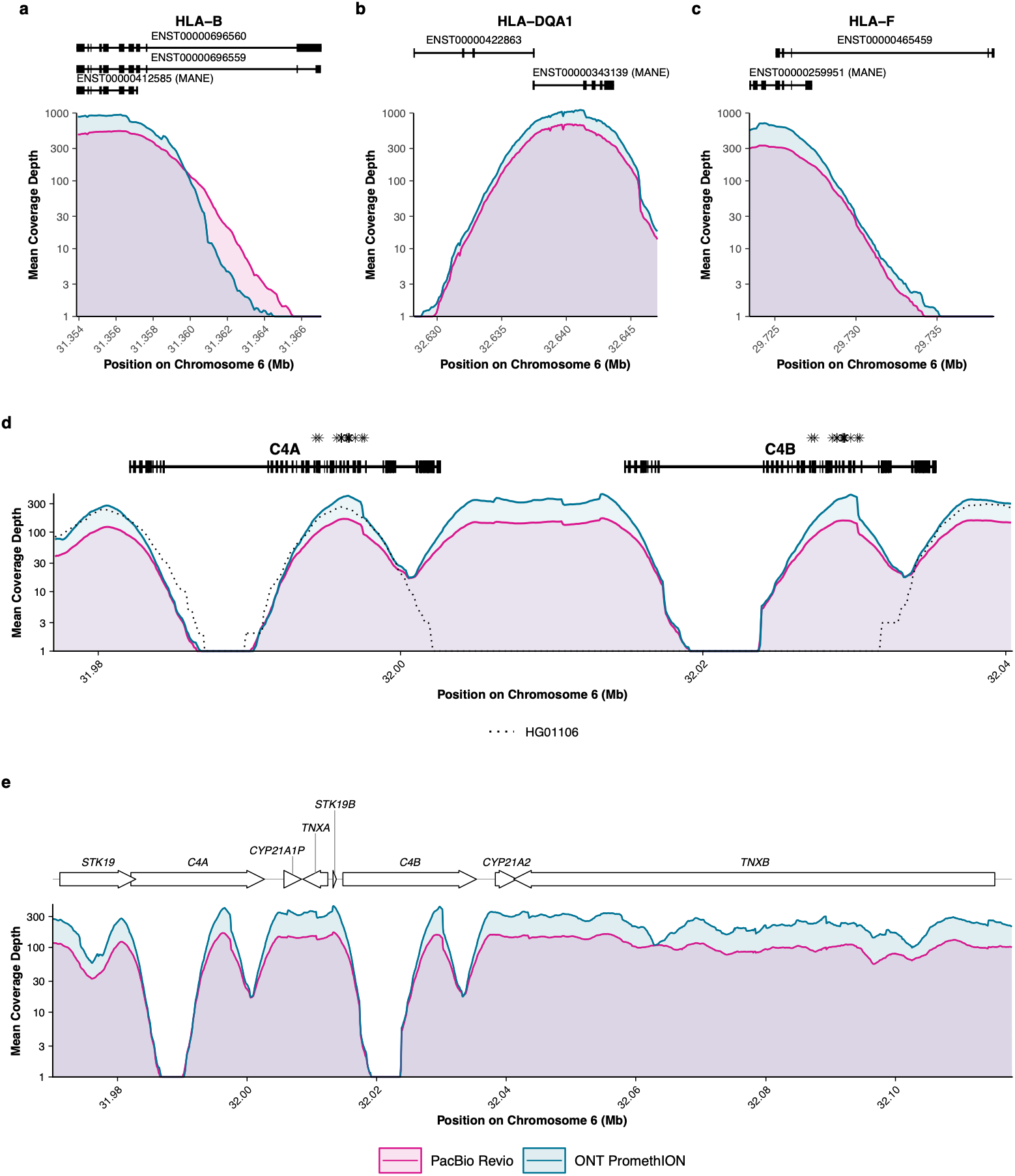
Coverage depth at loci with annotation adjustments or difficult mapping. Mean coverage depth (log scale) is shown for PacBio Revio (magenta) and ONT PromethION (teal), computed from per-base coverage averaged across samples and aggregated into non-overlapping 100 bp windows. (a) HLA-B with three annotated isoforms. (b) HLA-DQA1 with two annotated isoforms. (c) HLA-F with two annotated isoforms. (d) C4A and C4B, with nucleotide positions that differ between C4A and C4B in the GRCh38 human reference genome demarcated with asterisks. Coverage for HG01106 is shown as the mean depth in 100 bp windows (dotted line). (e) RCCX locus with genes annotated as labeled arrows. Coverage is capped at 1,000x for display in panels a-c. Because the y-axis is logarithmic, windows with mean depth below 1x are plotted at 1x.

## References

1. Kennedy, A.E., Ozbek, U., and Dorak, M.T. (2017). What has GWAS done for HLA and disease associations? Int. J. Immunogenet. 44, 195–211. 10.1111/iji.12332.

2. Sachidanandam, R., Weissman, D., Schmidt, S.C., Kakol, J.M., Stein, L.D., Marth, G., Sherry, S., Mullikin, J.C., Mortimore, B.J., Willey, D.L., et al. (2001). A map of human genome sequence variation containing 1.42 million single nucleotide polymorphisms. Nature 409, 928–933. 10.1038/35057149.

3. Durbin, R.M., Altshuler, D., Abecasis, G.R., Bentley, D.R., Chakravarti, A., Clark, A.G., Collins, F.S., De La Vega, F.M., Donnelly, P., Egholm, M., et al. (2010). A map of human genome variation from population-scale sequencing. Nature 467, 1061–1073. 10.1038/nature09534.

4. Long, X., and Xue, H. (2021). Genetic-variant hotspots and hotspot clusters in the human genome facilitating adaptation while increasing instability. Hum. Genomics 15, 19. 10.1186/s40246-021-00318-3.

5. Lenz, T.L., Spirin, V., Jordan, D.M., and Sunyaev, S.R. (2016). Excess of deleterious mutations around HLA genes reveals evolutionary cost of balancing selection. Mol. Biol. Evol. 33, 2555–2564. 10.1093/molbev/msw127.

6. Shiina, T., Hosomichi, K., Inoko, H., and Kulski, J.K. (2009). The HLA genomic loci map: expression, interaction, diversity and disease. J. Hum. Genet. 54, 15–39. 10.1038/jhg.2008.5.

7. Trowsdale, J., and Knight, J.C. (2013). Major histocompatibility complex genomics and human disease. Annu. Rev. Genomics Hum. Genet. 14, 301–323. 10.1146/annurev-genom-091212-153455.

8. Lee, S.J., Klein, J., Haagenson, M., Baxter-Lowe, L.A., Confer, D.L., Eapen, M., Fernandez-Vina, M., Flomenberg, N., Horowitz, M., Hurley, C.K., et al. (2007). High-resolution donor-recipient HLA matching contributes to the success of unrelated donor marrow transplantation. Blood 110, 4576–4583. 10.1182/blood-2007-06-097386.

9. Spellman, S.R., Eapen, M., Logan, B.R., Mueller, C., Rubinstein, P., Setterholm, M.I., Woolfrey, A.E., Horowitz, M.M., Confer, D.L., and Hurley, C.K. (2012). A perspective on the selection of unrelated donors and cord blood units for transplantation. Blood 120, 259–265. 10.1182/blood-2012-03-379032.

10. Montgomery, R.A., Tatapudi, V.S., Leffell, M.S., and Zachary, A.A. (2018). HLA in transplantation. Nat. Rev. Nephrol. 14, 558–570. 10.1038/s41581-018-0039-x.

11. Kok, G., Ilcken, E.F., Houwen, R.H.J., Lindemans, C.A., Nieuwenhuis, E.E.S., Spierings, E., and Fuchs, S.A. (2023). The effect of genetic HLA matching on liver transplantation outcome: A systematic review and meta-analysis. Ann. Surg. Open 4, e334. 10.1097/AS9.0000000000000334.

12. Cornaby, C., Schmitz, J.L., and Weimer, E.T. (2021). Next-generation sequencing and clinical histocompatibility testing. Hum. Immunol. 82, 829–837. 10.1016/j.humimm.2021.08.009.

13. Dilthey, A.T. (2021). State-of-the-art genome inference in the human MHC. Int. J. Biochem. Cell Biol. 131, 105882. 10.1016/j.biocel.2020.105882.

14. Brandt, D.Y.C., Aguiar, V.R.C., Bitarello, B.D., Nunes, K., Goudet, J., and Meyer, D. (2015). Mapping bias overestimates reference allele frequencies at the HLA genes in the 1000 Genomes Project Phase I data. G3 Genes Genomes Genet. 5, 931–941. 10.1534/g3.114.015784.

15. Castelli, E.C., Paz, M.A., Souza, A.S., Ramalho, J., and Mendes-Junior, C.T. (2018). *Hla-mapper*: An application to optimize the mapping of HLA sequences produced by massively parallel sequencing procedures. Hum. Immunol. 79, 678–684. 10.1016/j.humimm.2018.06.010.

16. Klasberg, S., Surendranath, V., Lange, V., and Schöfl, G. (2019). Bioinformatics strategies, challenges, and opportunities for next generation sequencing-based HLA genotyping. Transfus. Med. Hemotherapy 46, 312–325. 10.1159/000502487.

17. Bai, Y., Ni, M., Cooper, B., Wei, Y., and Fury, W. (2014). Inference of high resolution HLA types using genome-wide RNA or DNA sequencing reads. BMC Genomics 15, 325. 10.1186/1471-2164-15-325.

18. Szolek, A., Schubert, B., Mohr, C., Sturm, M., Feldhahn, M., and Kohlbacher, O. (2014). OptiType: precision HLA typing from next-generation sequencing data. Bioinformatics 30, 3310–3316. 10.1093/bioinformatics/btu548.

19. Shukla, S.A., Rooney, M.S., Rajasagi, M., Tiao, G., Dixon, P.M., Lawrence, M.S., Stevens, J., Lane, W.J., Dellagatta, J.L., Steelman, S., et al. (2015). Comprehensive analysis of cancer-associated somatic mutations in class I HLA genes. Nat. Biotechnol. 33, 1152–1158. 10.1038/nbt.3344.

20. Kawaguchi, S., Higasa, K., Shimizu, M., Yamada, R., and Matsuda, F. (2017). HLA-HD: An accurate HLA typing algorithm for next-generation sequencing data. Hum. Mutat. 38, 788–797. 10.1002/humu.23230.

21. Song, L., Bai, G., Liu, X.S., Li, B., and Li, H. (2023). Efficient and accurate KIR and HLA genotyping with massively parallel sequencing data. Genome Res. 33, 923–931. 10.1101/gr.277585.122.

22. Dilthey, A., Cox, C., Iqbal, Z., Nelson, M.R., and McVean, G. (2015). Improved genome inference in the MHC using a population reference graph. Nat. Genet. 47, 682–688. 10.1038/ng.3257.

23. Dilthey, A.T., Gourraud, P.-A., Mentzer, A.J., Cereb, N., Iqbal, Z., and McVean, G. (2016). High-accuracy HLA type inference from whole-genome sequencing data using population reference graphs. PLOS Comput. Biol. 12, e1005151. 10.1371/journal.pcbi.1005151.

24. Dilthey, A.T., Mentzer, A.J., Carapito, R., Cutland, C., Cereb, N., Madhi, S.A., Rhie, A., Koren, S., Bahram, S., McVean, G., et al. (2019). HLA*LA-HLA typing from linearly projected graph alignments. Bioinformatics 35, 4394–4396. 10.1093/bioinformatics/btz235.

25. Kim, D., Paggi, J.M., Park, C., Bennett, C., and Salzberg, S.L. (2019). Graph-based genome alignment and genotyping with HISAT2 and HISAT-genotype. Nat. Biotechnol. 37, 907–915. 10.1038/s41587-019-0201-4.

26. Liu, C., Yang, X., Duffy, B., Mohanakumar, T., Mitra, R.D., Zody, M.C., and Pfeifer, J.D. (2013). ATHLATES: accurate typing of human leukocyte antigen through exome sequencing. Nucleic Acids Res. 41, e142. 10.1093/nar/gkt481.

27. Huang, Y., Yang, J., Ying, D., Zhang, Y., Shotelersuk, V., Hirankarn, N., Sham, P.C., Lau, Y.L., and Yang, W. (2015). HLAreporter: a tool for HLA typing from next generation sequencing data. Genome Med. 7, 25. 10.1186/s13073-015-0145-3.

28. Lee, H., and Kingsford, C. (2018). Kourami: graph-guided assembly for novel human leukocyte antigen allele discovery. Genome Biol. 19, 16. 10.1186/s13059-018-1388-2.

29. Wang, S., Wang, M., Chen, L., Pan, G., Wang, Y., and Li, S.C. (2023). SpecHLA enables full-resolution HLA typing from sequencing data. Cell Rep. Methods 3, 100589. 10.1016/j.crmeth.2023.100589.

30. Duke, J.L., Mosbruger, T.L., Ferriola, D., Chitnis, N., Hu, T., Tairis, N., Margolis, D.J., and Monos, D.S. (2019). Resolving MiSeq-generated ambiguities in HLA-DPB1 typing by using the Oxford Nanopore Technology. J. Mol. Diagn. 21, 852–861. 10.1016/j.jmoldx.2019.04.009.

31. Centurião, N. de F., Maciel, G.C., Goldenstein, M.F.L., Alonso, E.O., and Torres, M.A. (2025). The novel HLA-C*03:678 allele identified using nanopore sequencing. HLA 105, e70038. 10.1111/tan.70038.

32. Allen, E.S., Yang, B., Garrett, J., Ball, E.D., Maiers, M., and Morris, G.P. (2018). Improved accuracy of clinical HLA genotyping by next-generation DNA sequencing affects unrelated donor search results for hematopoietic stem cell transplantation. Hum. Immunol. 79, 848–854. 10.1016/j.humimm.2018.10.008.

33. Madbouly, A., Gragert, L., Freeman, J., Leahy, N., Gourraud, P.-A., Hollenbach, J.A., Kamoun, M., Fernandez-Vina, M., and Maiers, M. (2014). Validation of statistical imputation of allele-level multilocus phased genotypes from ambiguous HLA assignments. Tissue Antigens 84, 285–292. 10.1111/tan.12390.

34. Maiers, M., Halagan, M., Gragert, L., Bashyal, P., Brelsford, J., Schneider, J., Lutsker, P., and Louzoun, Y. (2019). GRIMM: Graph imputation and matching for HLA genotypes. Bioinformatics 35, 3520–3523. 10.1093/bioinformatics/btz050.

35. Bochtler, W., Gragert, L., Patel, Z.I., Robinson, J., Steiner, D., Hofmann, J.A., Pingel, J., Baouz, A., Melis, A., Schneider, J., et al. (2016). A comparative reference study for the validation of HLA-matching algorithms in the search for allogeneic hematopoietic stem cell donors and cord blood units. HLA 87, 439–448. 10.1111/tan.12817.

36. Barker, D.J., Maccari, G., Georgiou, X., Cooper, M.A., Flicek, P., Robinson, J., and Marsh, S.G.E. (2023). The IPD-IMGT/HLA Database. Nucleic Acids Res. 51, D1053–D1060. 10.1093/nar/gkac1011.

37. Marsh, S.G.E., Osoegawa, K., Bodmer, W.F., Bontrop, R.E., Carrington, M.N., Erlich, H.A., Heidt, S., Holdsworth, R., Mayr, W.R., Maiers, M., et al. (2026). Nomenclature for factors of the HLA system, 2026. HLA 107, e70595. 10.1111/tan.70595.

38. Barker, D.J., Natarajan, R.H.L., Cooper, M.A., Hopper, S.J.F., Yates, A.D., Parham, P., Marsh, S.G.E., and Robinson, J. (2026). The IPD-IMGT/HLA database: recent developments in sequence submission. Nucleic Acids Res. 54, D1152–D1158. 10.1093/nar/gkaf1218.

39. Jain, M., Koren, S., Miga, K.H., Quick, J., Rand, A.C., Sasani, T.A., Tyson, J.R., Beggs, A.D., Dilthey, A.T., Fiddes, I.T., et al. (2018). Nanopore sequencing and assembly of a human genome with ultra-long reads. Nat. Biotechnol. 36, 338–345. 10.1038/nbt.4060.

40. Sedlazeck, F.J., Rescheneder, P., Smolka, M., Fang, H., Nattestad, M., von Haeseler, A., and Schatz, M.C. (2018). Accurate detection of complex structural variations using single-molecule sequencing. Nat. Methods 15, 461–468. 10.1038/s41592-018-0001-7.

41. Wenger, A.M., Peluso, P., Rowell, W.J., Chang, P.-C., Hall, R.J., Concepcion, G.T., Ebler, J., Fungtammasan, A., Kolesnikov, A., Olson, N.D., et al. (2019). Accurate circular consensus long-read sequencing improves variant detection and assembly of a human genome. Nat. Biotechnol. 37, 1155–1162. 10.1038/s41587-019-0217-9.

42. Logsdon, G.A., Vollger, M.R., and Eichler, E.E. (2020). Long-read human genome sequencing and its applications. Nat. Rev. Genet. 21, 597–614. 10.1038/s41576-020-0236-x.

43. Liao, W.-W., Asri, M., Ebler, J., Doerr, D., Haukness, M., Hickey, G., Lu, S., Lucas, J.K., Monlong, J., Abel, H.J., et al. (2023). A draft human pangenome reference. Nature 617, 312–324. 10.1038/s41586-023-05896-x.

44. Holt, J., Saunders, C.T., Rowell, W.J., Kronenberg, Z., Wenger, A.M., and Eberle, M. (2024). HiPhase: jointly phasing small, structural, and tandem repeat variants from HiFi sequencing. Bioinformatics 40, btae042. 10.1093/bioinformatics/btae042.

45. Liu, C., Xiao, F., Hoisington-Lopez, J., Lang, K., Quenzel, P., Duffy, B., and Mitra, R.D. (2018). Accurate typing of human leukocyte antigen class I genes by Oxford Nanopore sequencing. J. Mol. Diagn. 20, 428–435. 10.1016/j.jmoldx.2018.02.006.

46. Holt, J., Harting, J., Chen, X., Baker, D., Saunders, C.T., Kronenberg, Z., Gonzaludo, N., Yoo, B., Hudjashov, G., Jõeloo, M., et al. (2024). StarPhase: comprehensive phase-aware pharmacogenomic diplotyper for long-read sequencing data. Preprint at bioRxiv.

47. Wang, S., Wang, X., Wang, M., Zhou, Q., Wang, L., and Li, S.C. (2026). A scalable framework for comprehensive typing of polymorphic immune genes from long-read data. Adv. Sci. 13, e21531. 10.1002/advs.202521531.

48. Hu, J., Qin, Q., Li, H., and Zhou, Y. (2026). FuFiHLA: a tool for full-field HLA typing from long-read data. Bioinformatics 42, btag231. 10.1093/bioinformatics/btag231.

49. Mayor, N.P., Hayhurst, J.D., Turner, T.R., Szydlo, R.M., Shaw, B.E., Bultitude, W.P., Sayno, J.-R., Tavarozzi, F., Latham, K., Anthias, C., et al. (2019). Recipients receiving better HLA-matched hematopoietic cell transplantation grafts, uncovered by a novel HLA typing method, have superior survival: A retrospective study. Biol. Blood Marrow Transplant. 25, 443–450. 10.1016/j.bbmt.2018.12.768.

50. Mayor, N.P., Wang, T., Lee, S.J., Kuxhausen, M., Vierra-Green, C., Barker, D.J., Auletta, J., Bhatt, V.R., Gadalla, S.M., Gragert, L., et al. (2021). Impact of previously unrecognized HLA mismatches using ultrahigh resolution typing in unrelated donor hematopoietic cell transplantation. J. Clin. Oncol. 39, 2397–2409. 10.1200/JCO.20.03643.

51. Mayor, N.P., Hayhurst, J.D., Turner, T.R., Szydlo, R.M., Shaw, B.E., Bultitude, W.P., Sayno, J.-R., Tavarozzi, F., Latham, K., Anthias, C., et al. (2019). A reply to Hurley et al. regarding Recipients Receiving Better HLA-Matched Hematopoietic Cell Transplantation Grafts, Uncovered by a Novel HLA Typing Method, Have Superior Survival: A Retrospective Study. Biol. Blood Marrow Transplant. 25, e270–e271. 10.1016/j.bbmt.2019.06.005.

52. Hurley, C.K., Spellman, S., Dehn, J., Barker, J.N., Devine, S., Fernandez-Vina, M., Gautreaux, M., Logan, B., Maiers, M., Mueller, C., et al. (2019). Regarding “recipients receiving better HLA-matched hematopoietic cell transplantation grafts, uncovered by a novel HLA typing method, have superior survival: a retrospective study.” Biol. Blood Marrow Transplant. 25, e268–e269. 10.1016/j.bbmt.2019.05.026.

53. Handunnetthi, L., Ramagopalan, S.V., Ebers, G.C., and Knight, J.C. (2010). Regulation of major histocompatibility complex Class II gene expression, genetic variation and disease. Genes Immun. 11, 99–112. 10.1038/gene.2009.83.

54. Baxter-Lowe, L.A. (2021). The changing landscape of HLA typing: understanding how and when HLA typing data can be used with confidence from bench to bedside. Hum. Immunol. 82, 466–477. 10.1016/j.humimm.2021.04.011.

55. Arumugam, T., Adimulam, T., Gokul, A., and Ramsuran, V. (2024). Variation within the non-coding genome influences genetic and epigenetic regulation of the human leukocyte antigen genes. Front. Immunol. 15, 1422834. 10.3389/fimmu.2024.1422834.

56. Petersdorf, E.W., Malkki, M., O’hUigin, C., Carrington, M., Gooley, T., Haagenson, M.D., Horowitz, M.M., Spellman, S.R., Wang, T., and Stevenson, P. (2015). High HLA-DP Expression and Graft-versus-Host Disease. N. Engl. J. Med. 373, 599–609. 10.1056/NEJMoa1500140.

57. Sayer, D., Nytes, J., Jerkins, J.H., and Anderson, M.W. (2025). High rates of MHC mismatches in HLA matched unrelated donor/recipient pairs and potential impact on hematopoietic cell transplant outcome. Hum. Immunol. 86, 111186. 10.1016/j.humimm.2024.111186.

58. Carapito, R., Jung, N., Kwemou, M., Untrau, M., Michel, S., Pichot, A., Giacometti, G., Macquin, C., Ilias, W., Morlon, A., et al. (2016). Matching for the nonconventional MHC-I MICA gene significantly reduces the incidence of acute and chronic GVHD. Blood 128, 1979–1986. 10.1182/blood-2016-05-719070.

59. Clancy, J., Ritari, J., Lobier, M., Niittyvuopio, R., Salmenniemi, U., Putkonen, M., Itälä-Remes, M., Partanen, J., and Koskela, S. (2019). Increased MHC matching by *C4* gene compatibility in unrelated donor hematopoietic stem cell transplantation. Biol. Blood Marrow Transplant. 25, 891–898. 10.1016/j.bbmt.2018.12.759.

60. Petersdorf, E.W., McKallor, C., Malkki, M., Hsu, K., He, M., Spellman, S.R., Gooley, T., and Stevenson, P. (2025). The association of HLA-E ligand and NKG2 receptor variation with relapse and mortality after haploidentical related donor transplantation. Transplant. Cell. Ther. 31, 137–156. 10.1016/j.jtct.2025.01.004.

61. Yang, Y., Chung, E.K., Zhou, B., Lhotta, K., Hebert, L.A., Birmingham, D.J., Rovin, B.H., and Yu, Y. (2003). The intricate role of complement component C4 in human systemic lupus erythematosus. In Complement in Autoimmunity (Karger), pp. 98–132. 10.1159/000075689.

62. Schalkwijk, J., Zweers, M.C., Steijlen, P.M., Dean, W.B., Taylor, G., Vlijmen, I.M. van, Haren, B. van, Miller, W.L., and Bristow, J. (2001). A recessive form of the Ehlers–Danlos syndrome caused by tenascin-X deficiency. N. Engl. J. Med. 345, 1167–1175. 10.1056/NEJMoa002939.

63. Burch, G.H., Gong, Y., Liu, W., Dettman, R.W., Curry, C.J., Smith, L., Miller, W.L., and Bristow, J. (1997). Tenascin–X deficiency is associated with Ehlers–Danlos syndrome. Nat. Genet. 17, 104–108. 10.1038/ng0997-104.

64. White, P.C., and Speiser, P.W. (2000). Congenital adrenal hyperplasia due to 21-hydroxylase deficiency. Endocr. Rev. 21, 245–291. 10.1210/edrv.21.3.0398.

65. Monlong, J., Chen, X., Barseghyan, H., Rowell, W.J., Negi, S., Nokoff, N., Mohnach, L., Hirsch, J., Finlayson, C., Keegan, C.E., et al. (2025). Long-read sequencing resolves the clinically relevant CYP21A2 locus, supporting a new clinical test for Congenital Adrenal Hyperplasia. Preprint at medRxiv.

66. Wade, K.J., Suseno, R., Kizer, K., Williams, J., Boquett, J., Caillier, S., Pollock, N.R., Renschen, A., Santaniello, A., Oksenberg, J.R., et al. (2024). MHConstructor: a high-throughput, haplotype-informed solution to the MHC assembly challenge. Genome Biol. 25, 274. 10.1186/s13059-024-03412-6.

67. Marin, W.M., Augusto, D.G., Wade, K.J., and Hollenbach, J.A. (2024). High-throughput complement component 4 genomic sequence analysis with C4Investigator. HLA 103, e15273. 10.1111/tan.15273.

68. Albrecht, V., Zweiniger, C., Surendranath, V., Lang, K., Schöfl, G., Dahl, A., Winkler, S., Lange, V., Böhme, I., and Schmidt, A.H. (2017). Dual redundant sequencing strategy: full-length gene characterisation of 1056 novel and confirmatory HLA alleles. HLA 90, 79–87. 10.1111/tan.13057.

69. Liu, C., Yang, X., Duffy, B.F., Hoisington-Lopez, J., Crosby, M., Porche-Sorbet, R., Saito, K., Berry, R., Swamidass, V., and Mitra, R.D. (2021). High-resolution HLA typing by long reads from the R10.3 Oxford nanopore flow cells. Hum. Immunol. 82, 288–295. 10.1016/j.humimm.2021.02.005.

70. Stockton, J.D., Nieto, T., Wroe, E., Poles, A., Inston, N., Briggs, D., and Beggs, A.D. (2020). Rapid, highly accurate and cost-effective open-source simultaneous complete HLA typing and phasing of class I and II alleles using nanopore sequencing. HLA 96, 163–178. 10.1111/tan.13926.

71. El-Lagta, N., Truong, L., Ayora, F., Mobegi, F., Bruce, S., Martinez, P., D’Orsogna, L., and De Santis, D. (2024). Revolutionising high resolution HLA genotyping for transplant assessment: validation, implementation and challenges of oxford nanopore technologies’ Q20+ sequencing. HLA 104, e15725. 10.1111/tan.15725.

72. Yin, Y., Lan, J.H., Nguyen, D., Valenzuela, N., Takemura, P., Bolon, Y.-T., Springer, B., Saito, K., Zheng, Y., Hague, T., et al. (2016). Application of high-throughput next-generation sequencing for HLA typing on buccal extracted DNA: results from over 10,000 donor recruitment samples. PLOS ONE 11, e0165810. 10.1371/journal.pone.0165810.

73. Laver, T.W., Caswell, R.C., Moore, K.A., Poschmann, J., Johnson, M.B., Owens, M.M., Ellard, S., Paszkiewicz, K.H., and Weedon, M.N. (2016). Pitfalls of haplotype phasing from amplicon-based long-read sequencing. Sci. Rep. 6, 21746. 10.1038/srep21746.

74. van Deutekom, H.W.M., Mulder, W., and Rozemuller, E.H. (2019). Accuracy of NGS HLA typing data influenced by STR. Hum. Immunol. 80, 461–464. 10.1016/j.humimm.2019.03.007.

75. Steiert, T.A., Fuß, J., Juzenas, S., Wittig, M., Hoeppner, M.P., Vollstedt, M., Varkalaite, G., ElAbd, H., Brockmann, C., Görg, S., et al. (2022). High-throughput method for the hybridisation-based targeted enrichment of long genomic fragments for PacBio third-generation sequencing. NAR Genomics Bioinforma. 4, lqac051. 10.1093/nargab/lqac051.

76. Wittig, M., Anmarkrud, J.A., Kässens, J.C., Koch, S., Forster, M., Ellinghaus, E., Hov, J.R., Sauer, S., Schimmler, M., Ziemann, M., et al. (2015). Development of a high-resolution NGS-based HLA-typing and analysis pipeline. Nucleic Acids Res. 43, e70. 10.1093/nar/gkv184.

77. Norman, P.J., Norberg, S.J., Guethlein, L.A., Nemat-Gorgani, N., Royce, T., Wroblewski, E.E., Dunn, T., Mann, T., Alicata, C., Hollenbach, J.A., et al. (2017). Sequences of 95 human MHC haplotypes reveal extreme coding variation in genes other than highly polymorphic HLA class I and II. Genome Res. 27, 813–823. 10.1101/gr.213538.116.

78. Lai, S.-K., Luo, A.C., Chiu, I.-H., Chuang, H.-W., Chou, T.-H., Hung, T.-K., Hsu, J.S., Chen, C.-Y., Yang, W.-S., Yang, Y.-C., et al. (2024). A novel framework for human leukocyte antigen (HLA) genotyping using probe capture-based targeted next-generation sequencing and computational analysis. Comput. Struct. Biotechnol. J. 23, 1562–1571. 10.1016/j.csbj.2024.03.030.

79. Robinson, J.T., Thorvaldsdóttir, H., Winckler, W., Guttman, M., Lander, E.S., Getz, G., and Mesirov, J.P. (2011). Integrative genomics viewer. Nat. Biotechnol. 29, 24–26. 10.1038/nbt.1754.

80. Krusche, P., Trigg, L., Boutros, P.C., Mason, C.E., De La Vega, F.M., Moore, B.L., Gonzalez-Porta, M., Eberle, M.A., Tezak, Z., Lababidi, S., et al. (2019). Best practices for benchmarking germline small-variant calls in human genomes. Nat. Biotechnol. 37, 555–560. 10.1038/s41587-019-0054-x.

81. Chen, X., Harting, J., Farrow, E., Thiffault, I., Kasperaviciute, D., Hoischen, A., Gilissen, C., Pastinen, T., and Eberle, M.A. (2023). Comprehensive *SMN1* and *SMN2* profiling for spinal muscular atrophy analysis using long-read PacBio HiFi sequencing. Am. J. Hum. Genet. 110, 240–250. 10.1016/j.ajhg.2023.01.001.

82. Satta, Y., Mayer, W.E., and Klein, J. (1996). HLA-DRB intron 1 sequences: implications for the evolution of HLA-DRB genes and haplotypes. Hum. Immunol. 51, 1–12. 10.1016/S0198-8859(96)00155-3.

83. Gustafson, J.A., Gibson, S.B., Damaraju, N., Zalusky, M.P.G., Hoekzema, K., Twesigomwe, D., Yang, L., Snead, A.A., Richmond, P.A., De Coster, W., et al. (2024). High-coverage nanopore sequencing of samples from the 1000 Genomes Project to build a comprehensive catalog of human genetic variation. Genome Res. 34, 2061–2073. 10.1101/gr.279273.124.

84. Osoegawa, K., Vayntrub, T.A., Wenda, S., De Santis, D., Barsakis, K., Ivanova, M., Hsu, S., Barone, J., Holdsworth, R., Diviney, M., et al. (2019). Quality control project of NGS HLA genotyping for the 17th International HLA and Immunogenetics Workshop. Hum. Immunol. 80, 228–236. 10.1016/j.humimm.2019.01.009.

85. Creary, L.E., Guerra, S.G., Chong, W., Brown, C.J., Turner, T.R., Robinson, J., Bultitude, W.P., Mayor, N.P., Marsh, S.G.E., Saito, K., et al. (2019). Next-generation HLA typing of 382 International Histocompatibility Working Group reference B-lymphoblastoid cell lines: Report from the 17th International HLA and Immunogenetics Workshop. Hum. Immunol. 80, 449–460. 10.1016/j.humimm.2019.03.001.

86. Thuesen, N.H., Klausen, M.S., Gopalakrishnan, S., Trolle, T., and Renaud, G. (2022). Benchmarking freely available HLA typing algorithms across varying genes, coverages and typing resolutions. Front. Immunol. 13, 987655. 10.3389/fimmu.2022.987655.

87. Abi-Rached, L., Gouret, P., Yeh, J.-H., Di Cristofaro, J., Pontarotti, P., Picard, C., and Paganini, J. (2018). Immune diversity sheds light on missing variation in worldwide genetic diversity panels. PLOS ONE 13, e0206512. 10.1371/journal.pone.0206512.

88. Zhou, Y., Song, L., and Li, H. (2024). Full resolution HLA and KIR gene annotations for human genome assemblies. Genome Res. 34, 1931–1941. 10.1101/gr.278985.124.

89. Pappas, D.J., Lizee, A., Paunic, V., Beutner, K.R., Motyer, A., Vukcevic, D., Leslie, S., Biesiada, J., Meller, J., Taylor, K.D., et al. (2018). Significant variation between SNP-based HLA imputations in diverse populations: the last mile is the hardest. Pharmacogenomics J. 18, 367–376. 10.1038/tpj.2017.7.

90. Martin, M. (2011). Cutadapt removes adapter sequences from high-throughput sequencing reads. EMBnet.j. 17, 10–12. 10.14806/ej.17.1.200.

91. Li, H. (2018). Minimap2: Pairwise alignment for nucleotide sequences. Bioinformatics 34, 3094–3100. 10.1093/bioinformatics/bty191.

92. Broad Institute (2019). Picard Toolkit. (Broad Institute).

93. Pedersen, B.S., and Quinlan, A.R. (2018). Mosdepth: quick coverage calculation for genomes and exomes. Bioinformatics 34, 867–868. 10.1093/bioinformatics/btx699.

94. Poplin, R., Chang, P.-C., Alexander, D., Schwartz, S., Colthurst, T., Ku, A., Newburger, D., Dijamco, J., Nguyen, N., Afshar, P.T., et al. (2018). A universal SNP and small-indel variant caller using deep neural networks. Nat. Biotechnol. 36, 983–987. 10.1038/nbt.4235.

95. Zheng, Z., Li, S., Su, J., Leung, A.W.-S., Lam, T.-W., and Luo, R. (2022). Symphonizing pileup and full-alignment for deep learning-based long-read variant calling. Nat. Comput. Sci. 2, 797–803. 10.1038/s43588-022-00387-x.

96. Smolka, M., Paulin, L.F., Grochowski, C.M., Horner, D.W., Mahmoud, M., Behera, S., Kalef-Ezra, E., Gandhi, M., Hong, K., Pehlivan, D., et al. (2024). Detection of mosaic and population-level structural variants with Sniffles2. Nat. Biotechnol. 42, 1571–1580. 10.1038/s41587-023-02024-y.

97. Dolzhenko, E., English, A., Dashnow, H., De Sena Brandine, G., Mokveld, T., Rowell, W.J., Karniski, C., Kronenberg, Z., Danzi, M.C., Cheung, W.A., et al. (2024). Characterization and visualization of tandem repeats at genome scale. Nat. Biotechnol. 42, 1606–1614. 10.1038/s41587-023-02057-3.

98. Lin, J.-H., Chen, L.-C., Yu, S.-C., and Huang, Y.-T. (2022). LongPhase: an ultra-fast chromosome-scale phasing algorithm for small and large variants. Bioinformatics 38, 1816–1822. 10.1093/bioinformatics/btac058.

99. Patterson, M., Marschall, T., Pisanti, N., van Iersel, L., Stougie, L., Klau, G.W., and Schönhuth, A. (2015). WhatsHap: Weighted haplotype assembly for future-generation sequencing reads. J. Comput. Biol. 22, 498–509. 10.1089/cmb.2014.0157.

100. Cleary, J.G., Braithwaite, R., Gaastra, K., Hilbush, B.S., Inglis, S., Irvine, S.A., Jackson, A., Littin, R., Rathod, M., Ware, D., et al. (2015). Comparing variant call files for performance benchmarking of next-generation sequencing variant calling pipelines. Preprint at bioRxiv, 10.1101/023754 https://doi.org/10.1101/023754.

101. Chen, S. (2023). Ultrafast one-pass FASTQ data preprocessing, quality control, and deduplication using fastp. iMeta 2, e107. 10.1002/imt2.107.

102. Wang, J.R., and Li, H. (2026). Memory-safe high-performance sequence mapping with rammap. Preprint at bioRxiv.

103. Li, H. (2011). A statistical framework for SNP calling, mutation discovery, association mapping and population genetical parameter estimation from sequencing data. Bioinformatics 27, 2987–2993. 10.1093/bioinformatics/btr509.

104. Sanchez-Ramirez, S. (2017). vcf2fasta. https://github.com/santiagosnchez/vcf2fasta https://doi.org/https://github.com/santiagosnchez/vcf2fasta.

105. Šošić, M., and Šikić, M. (2017). Edlib: a C/C++ library for fast, exact sequence alignment using edit distance. Bioinformatics 33, 1394–1395. 10.1093/bioinformatics/btw753.

106. Danecek, P., Bonfield, J.K., Liddle, J., Marshall, J., Ohan, V., Pollard, M.O., Whitwham, A., Keane, T., McCarthy, S.A., Davies, R.M., et al. (2021). Twelve years of SAMtools and BCFtools. GigaScience 10, giab008. 10.1093/gigascience/giab008.

107. Quinlan, A.R., and Hall, I.M. (2010). BEDTools: a flexible suite of utilities for comparing genomic features. Bioinformatics 26, 841–842. 10.1093/bioinformatics/btq033.

108. Alexandrov, N., Wang, T., Blair, L., Nadon, B., and Sayer, D. (2023). HLA-OLI: a new MHC class I pseudogene and HLA-Y are located on a 60 kb indel in the human MHC between HLA-W and HLA-J. HLA 102, 599–606. 10.1111/tan.15180.

